# APOE genotype and the effect of statins: a systematic review and meta-analysis

**DOI:** 10.1101/2024.12.13.24318973

**Authors:** Innocent G. Asiimwe, Tsegay Gebru, Andrea L. Jorgensen, Munir Pirmohamed, Multimorbidity Mechanism and Therapeutic Research Collaborative (MMTRC)

**Affiliations:** Department of Pharmacology and Therapeutics, Institute of Systems, Molecular and Integrative Biology, University of Liverpool, Liverpool, UK; Department of Health Data Science, Institute of Population Health Sciences, University of Liverpool, Liverpool, UK

**Keywords:** *APOE* genotype, Lipid response, Meta-analysis, Statins, Systematic review

## Abstract

**Introduction:** The *APOE* genotype may affect statin therapy response. We conducted a systematic review and meta-analysis to update and quantify this association across various outcomes.

**Methods:** We searched seven databases (MEDLINE, Scopus, Web of Science, The Cochrane Library, APA PsycINFO, CINAHL Plus, and ClinicalTrials.gov) on 9^th^ May 2024. Screening and data extraction were performed by two reviewers and a machine learning tool (ASReview).

**Results:** From 4,352 de-duplicated records, 68 studies were included in the systematic review and 52 in the meta-analysis. Biomarkers analysed included Low-Density Lipoprotein Cholesterol (LDLC), Total Cholesterol (TC), Triglycerides (TG), and High-Density Lipoprotein Cholesterol (HDLC). Compared to *ε3* carriers, *ε2* carriers showed greater reductions in LDLC (mean difference: −2.98%, 95% CI: −5.88% to −0.08%) and similar reductions in TC (−2.73%, −5.62% to 0.16%), and TG (−4.95%, −11.93% to 2.04%) with no significant difference in HDLC (−0.09%, −3.10% to 2.91%). After adjusting for publication bias, *ε4* carriers showed less pronounced statin effects, with smaller reductions in LDLC (mean difference: 10.04%, 6.04% to 14.04%), TC (8.99%, 5.08% to 12.90%), and TG (8.24%, 2.15% to 14.33%), along with a smaller increase in HDLC (−10.08%, −15.30% to −4.85%) compared to *ε3* carriers. Study quality was uncertain, and heterogeneity (partly explained by sex and Familial Hypercholesterolemia) was high, especially for the percentage changes. A stronger genotype effect was seen in males.

**Conclusion:** Our meta-analysis shows that *APOE* genotype can significantly influence statin response, emphasizing the need to incorporate known genetic factors into personalized treatment regimes.

## Introduction

Cardiovascular diseases (CVD) remain the leading cause of death worldwide. In 2021, despite the COVID- 19 pandemic, ischaemic heart disease was the leading cause of age-standardised deaths globally, with 108.7 deaths (95% uncertainty interval [UI] 99.8 to 115.6) per 100,000 population.(1) Stroke, overtaken by COVID-19, ranked third with 87.4 deaths (95% UI 79.5 to 94.4) per 100,000 population.(1) Randomized controlled trials (RCTs) have consistently demonstrated the efficacy of statins, or 3-hydroxymethyl-3- methylglutaryl coenzyme A (HMG-CoA) reductase inhibitors, in reducing overall mortality, establishing them as standard treatments for primary and secondary prevention of cardiovascular disease. For example, a 2022 systematic review evaluating statin use for the primary prevention of CVD reported that statins significantly reduced the risk of all-cause mortality (risk ratio [RR] 0.92 [95% confidence interval/CI, 0.87 to 0.98]).(2) This review included evidence from 22 RCTs with a total of 90,624 participants. Statins were also protective against CVD, reducing the risk of fatal or nonfatal stroke (RR 0.78 [95% CI 0.68 to 0.90]) and myocardial infarction (RR 0.67 [95% CI 0.60 to 0.75]).(2)

The Apolipoprotein E (*APOE*) gene, located on the long arm of chromosome 19 (19q13.32), is both a CVD risk factor and a modulator of statin therapy.(3–5) It encodes the Apolipoprotein E (Apo E) protein, which plays a crucial role in lipid metabolism and is present in triglyceride-rich chylomicrons, very-low-density lipoproteins (VLDL), intermediate-density lipoproteins (IDL), and certain high-density lipoprotein (HDL) subclasses. There are three primary isoforms of Apo E (*ε2*, *ε3*, and *ε4*) resulting from two single-nucleotide polymorphisms (SNPs), rs429358 (T>C) and rs7412 (C>T). These SNPs result in key amino acid changes; specifically, rs429358 causes a cysteine (amino acid codon TGC) to arginine (CGC) change at position 112, and rs7412 causes an arginine to cysteine change at position 158. The *ε2* isoform has cysteine residues at both positions 112 and 158, the *ε3* isoform has cysteine at 112 and arginine at 158, and the *ε4* isoform has arginine at both positions.(6–8) The *ε3* allele is the most common, found in over 60% of the population.(3, 7) These three alleles can form six genotypes including three homozygotes (*ε2ε2*, *ε3ε3*, *ε4ε4*) and three heterozygotes (*ε2ε3*, *ε2ε4*, *ε3ε4*).

Apo E isoforms affect the metabolism and clearance of lipoproteins through interactions with receptors such as the low-density lipoprotein (LDL) receptor (LDLR).(6, 8, 9) The *ε2* isoform has defective binding to LDLR leading to delayed clearance, higher plasma Apo E levels, and upregulation of HMG-CoA and LDLR synthesis, resulting in lower plasma total cholesterol (TC) and LDL cholesterol (LDLC) levels. Conversely, the *ε4* isoform is cleared more rapidly, causing downregulation of HMG-CoA and LDLR, and consequently higher plasma TC and LDLC levels. This translates to *ε4* being associated with a higher risk of cardiovascular and neurological diseases, such as Alzheimer’s disease, whereas *ε2* is associated with a lower risk.(6, 9) For example, Bennet and colleagues analysed data from 82 studies on lipid levels (involving 86,067 healthy participants) and 121 studies on coronary outcomes (including 37,850 cases and 82,727 controls).(10) They found a consistent relationship between *APOE* genotypes, LDLC levels, and coronary risk. Compared to individuals with the *ε3ε3* genotype, *ε2* carriers had a 20% lower risk of coronary heart disease, while *ε4* carriers showed a slightly elevated risk. However, in rare cases, individuals with two ε2 alleles may develop familial dysbetalipoproteinemia, a condition that increases cardiovascular disease risk due to abnormal lipid metabolism.(3, 11)

As stated earlier, *APOE* genotype can influence responses to statin therapy, with *ε2* carriers generally experiencing more significant reductions in LDLC levels compared to *ε4* carriers.(8) However, if baseline LDLC levels are substantially higher in ε4 carriers, they may show greater percentage reductions in response to statin therapy when compared to *ε2* or *ε3* carriers who start with lower baseline levels.(12) Understanding the impact of *APOE* genotype on lipid metabolism and statin efficacy is crucial for managing cardiovascular risk and tailoring treatment strategies. Previous studies examining the role of *APOE* genotype in statin therapy response have produced inconsistent findings. For example, a 2009 systematic review analysed 24 studies and found no significant difference in the pooled mean reduction of total cholesterol among the genotypes: *ε2* carriers had a reduction of −27.7% (95% CI: −32.5 to −22.8%), *ε3ε3* had −25.3% (95% CI: −28.0 to −22.6%), and *ε4* carriers had −25.1% (95% CI: −29.3 to −21.0%), which translates to mean differences of −2.4% between *ε2* and *ε3* carriers and 0.2% between *ε4* and *ε3* carriers.(13) Similarly, there were no significant differences in LDLC, HDL cholesterol, or triglyceride levels across the genotype groups. To provide a comprehensive update on these findings, our systematic review aimed to quantify the association between *APOE* genotype and responses to statin therapy, including changes in lipid levels.

## Methods

This study followed a predefined protocol (PROSPERO: CRD42024545603) and is reported as per the Preferred Reporting Items for Systematic Reviews and Meta-Analyses (PRISMA) 2020 statement (Table S1).(14)

### Search Strategy and Selection Criteria

We searched seven databases (MEDLINE, Scopus, Web of Science, The Cochrane Library, APA PsycInfo, CINAHL Plus, and ClinicalTrials.gov) on 9^th^ May 2024 using medical subject headings and keywords related to “*APOE*” and “statins” (detailed search strategies are provided in Table S2). Additionally, we manually searched reference lists from identified studies and previous systematic reviews, and we contacted experts to identify further eligible articles.

We included all studies regardless of their publication year or status. Both observational studies (e.g., retrospective or prospective cohort and case-control studies) and interventional studies (e.g., randomized controlled trials) were considered if they investigated the association between (a) Apolipoprotein E (*APOE*) genotype (*APOE* SNPs such as rs429358 and rs7412 and *APOE* carrier status), (b) statins (including atorvastatin, cerivastatin, fluvastatin, lovastatin, pitavastatin, pravastatin, and simvastatin), and (c) any clinical outcomes related to safety and efficacy, including lipid levels, reported in the primary papers. We excluded case reports, review articles, letters, commentaries, and editorials unless they contained information on primary studies not published elsewhere. Non-English studies without translation and any studies from which data could not be extracted were also excluded.

### Data Extraction and Quality Assessment

The screening of titles, abstracts, and full texts of all retrieved bibliographic records was conducted by two reviewers: IGA (who screened all records) and TG (who screened MEDLINE records), along with a machine learning framework, ASReview.(15) ASReview screened all records, with stopping criteria based on the number of records selected by the human reviewers. If an abstract was unavailable, the full text was obtained unless the article could be confidently excluded based on its title alone. In cases of uncertainty regarding a study’s eligibility, it proceeded to the full text screening stage to minimize the risk of erroneously excluding relevant studies.

We created and piloted a data extraction form using a randomly selected subset of included papers to capture pertinent details such as study design, patient characteristics, study quality, and outcomes. When multiple studies analysed the same dataset (identified based on study acronyms, recruitment sites and periods, and authors and their affiliations) for a specific exposure-outcome combination, we prioritized peer-reviewed publications and those reporting data from a larger number of patients to avoid duplicate participant inclusion. We used WebPlotDigitizer (version 4, https://apps.automeris.io/wpd4/) to digitize and extract data (both central tendency and variability measures) presented only in figures.

To assess the methodological quality of each included study, we planned to use the checklist provided by the STrengthening the Reporting Of Pharmacogenetic Studies (STROPS) guideline.(16)

### Data Synthesis and Analysis

When two or more studies were available for variant-outcome combinations, we obtained pooled estimates by conducting pairwise meta-analyses comparing heterozygotes versus wild-type homozygotes, mutant-type homozygotes versus wild-type homozygotes, and/or mutant-type homozygotes plus heterozygotes versus wild-type homozygotes. For *APOE* carrier status, we conducted the following analyses: *ε2* carriers versus *ε3* carriers, *ε4* carriers versus *ε3* carriers, *ε2* carriers versus *ε2* non-carriers, and *ε4* carriers versus *ε4* non-carriers. In this context, *ε2* carriers included *ε2ε2* and *ε2ε3*, *ε3* carriers included *ε3ε3*, and *ε4* carriers included *ε3ε4* and *ε4ε4*. Due to the opposing effects of the *ε2* and *ε4* alleles, individuals with the *ε2ε4* genotype were initially excluded. However, sensitivity analyses were conducted, including *ε2ε4* individuals classified as *ε2*, *ε3*, or *ε4* carriers.

We considered outcomes recorded at baseline, post-treatment, or as changes from baseline separately. In each analysis, we ensured uniform units and used conversion factors reported in the literature. For example, to convert total cholesterol, low-density lipoprotein cholesterol, and high-density lipoprotein cholesterol from mg/dL to mmol/L, we divided the value in mg/dL by 38.67. To convert Apolipoprotein B from mg/dL to mmol/L, we assumed that 0.0512 mg/dL equals 1,000,000 mmol/L.(17)

Meta-analyses were performed using the meta(18) package in R (version 4.4.0). We generated odds ratios for dichotomous outcomes and mean differences for continuous outcomes, together with corresponding 95% confidence intervals. For studies reporting odds ratios, we pooled the reported estimates, giving priority to the estimates adjusting for the most covariates when multiple estimates were available. For continuous outcomes, we estimated means and standard deviations from provided median values and interquartile ranges,(19, 20) and combined means and standard deviations using formulas from the Cochrane Handbook(20) (Table S3) when necessary. Forest plots were created for the exposure-outcome combinations to visually represent the results.

We evaluated the magnitude of inconsistency in study results through visual inspection of forest plots and by considering the *I²* statistic,(20) categorizing heterogeneity as low (*I²* < 30%), moderate (30-70%), or high (> 70%). Potential sources of heterogeneity were explored in subgroup analyses based on factors such as sex, and Familial Hypercholesterolemia.

When ten or more studies were available for a given exposure-outcome combination, we assessed publication bias using the linear regression test of funnel plot asymmetry. A p-value < 0.1 was considered indicative of publication bias. If visual assessment suggested publication bias, we conducted exploratory analyses and adjusted for it using trim and fill analysis.

## Results

### Studies included in the Systematic Review

Figure 1 shows the literature search and selection process. From 4,352 unique records identified, 68 were included in the systematic review. The characteristics of these studies are detailed in Table S4, while Tables S5–S7 present the extracted results for the ratio, binary and continuous outcomes, respectively. The median publication year of these studies was 2008, with an interquartile range from 2002 to 2018.

**Figure 1.**
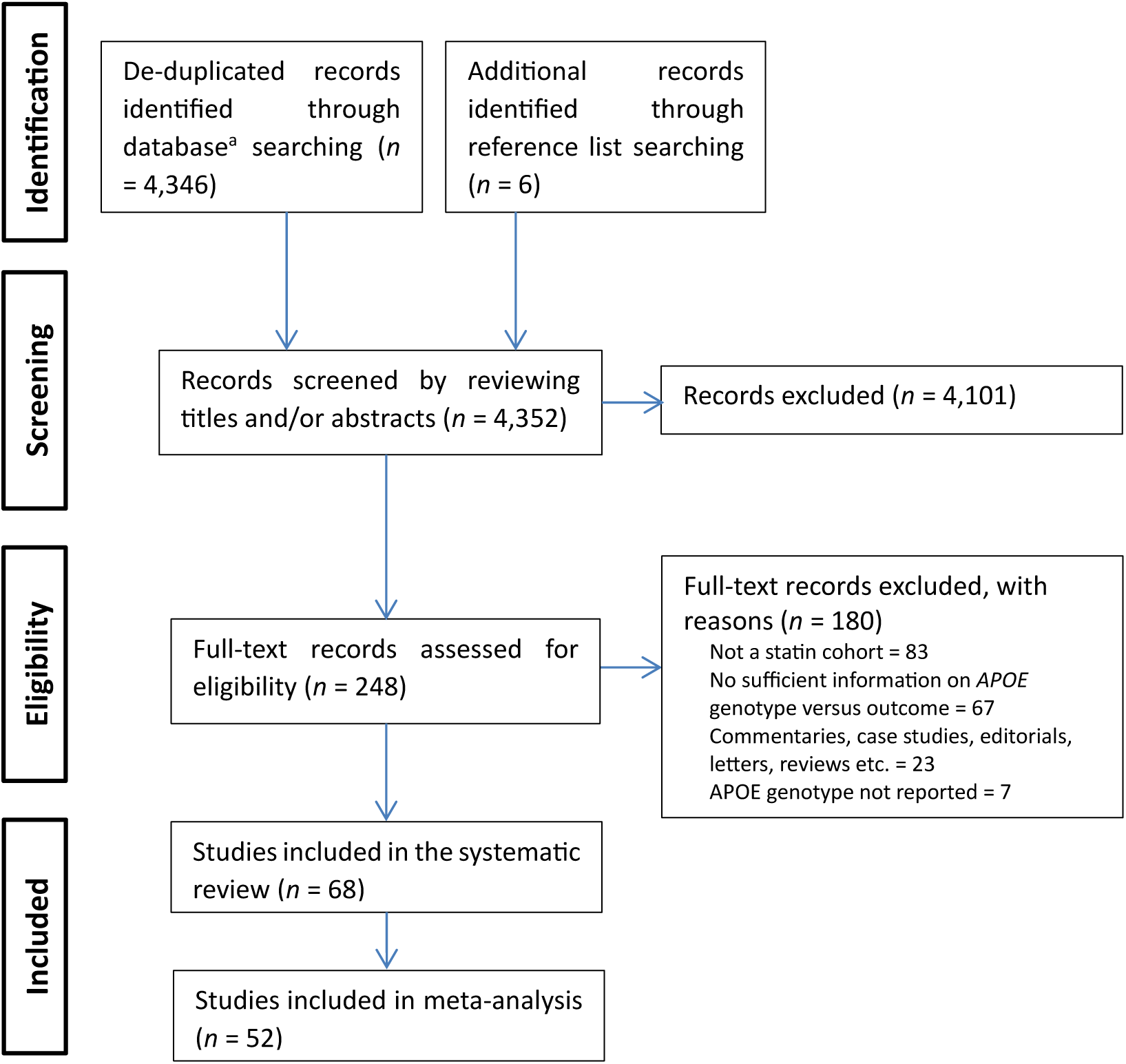
Flow chart of included studies. ^a^Databases included MEDLINE, Scopus, Web of Science, The Cochrane Library, APA PsycInfo, CINAHL Plus, and ClinicalTrials.gov. *APOE* = Apolipoprotein E.

We initially planned to assess the methodological quality of the included studies using the STROPS checklist. However, given that more than half of these studies (56%, or 38 out of 68) were published before 2010 (with 15%, or 10 out of 68, published before 2000), many key items—such as the Reference SNP cluster ID, sample size, genotype quality control methods, and population stratification—were not mandatory at that time. Based on a randomly selected subset of included papers, and due to the frequent lack of required information, we decided not to assess the methodological quality.

### Studies and Outcomes included in Meta-analysis

Of the 68 studies included in the systematic review, 52 were included in the meta-analysis. This included one study that reported adjusted odds ratios for two cohorts (Risk of lobar intracranial haemorrhage outcome),(21) two studies that reported mortality as a binary outcome,(22, 23) and 49 studies that reported continuous outcomes (Figure 2). Figure 2 is stratified by the time of biomarker measurement and includes 31 studies that reported biomarker measurements before statin treatment, 22 studies that reported measurements after statin treatment, and 35 studies that reported changes in biomarker levels.

**Figure 2.**
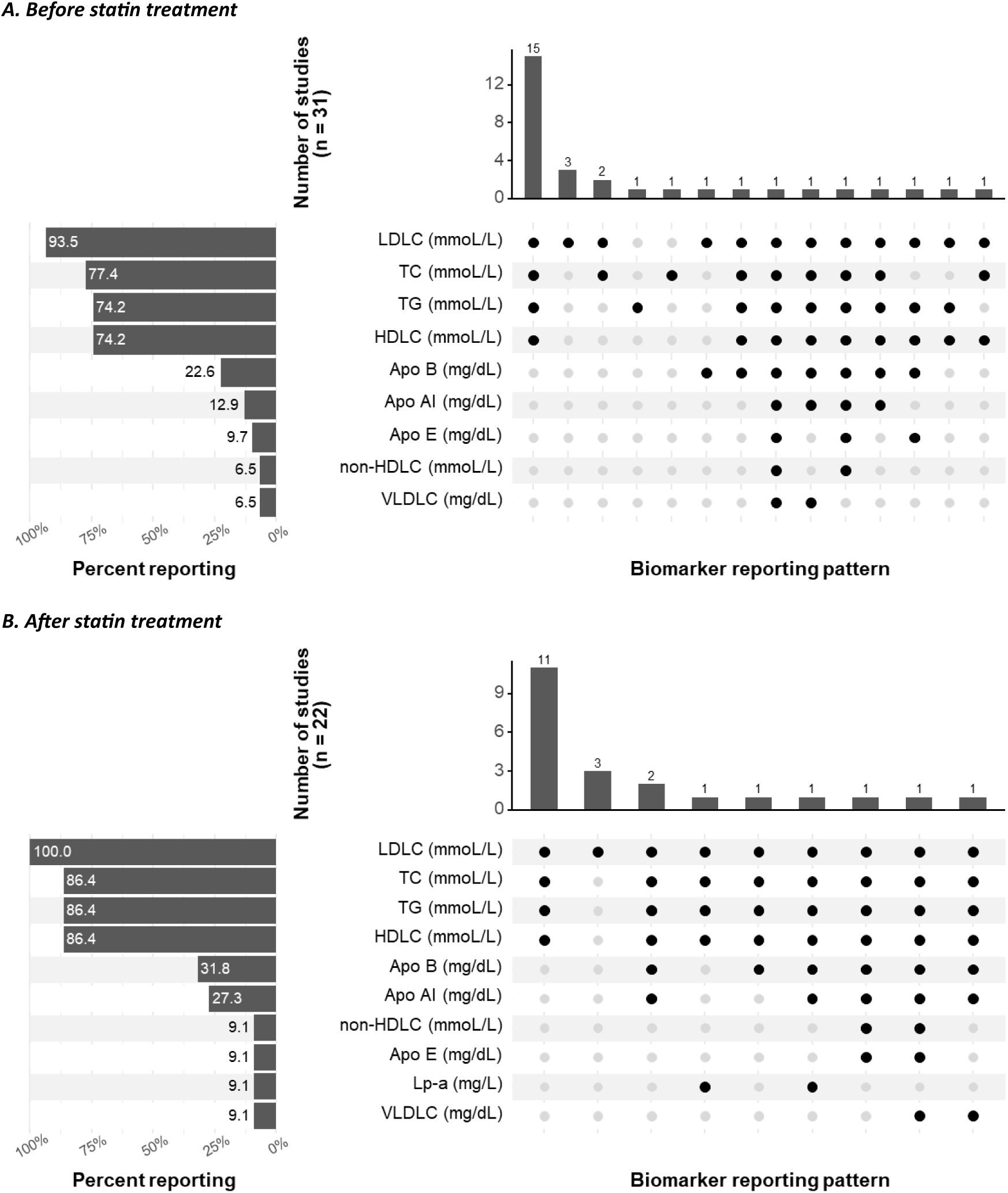
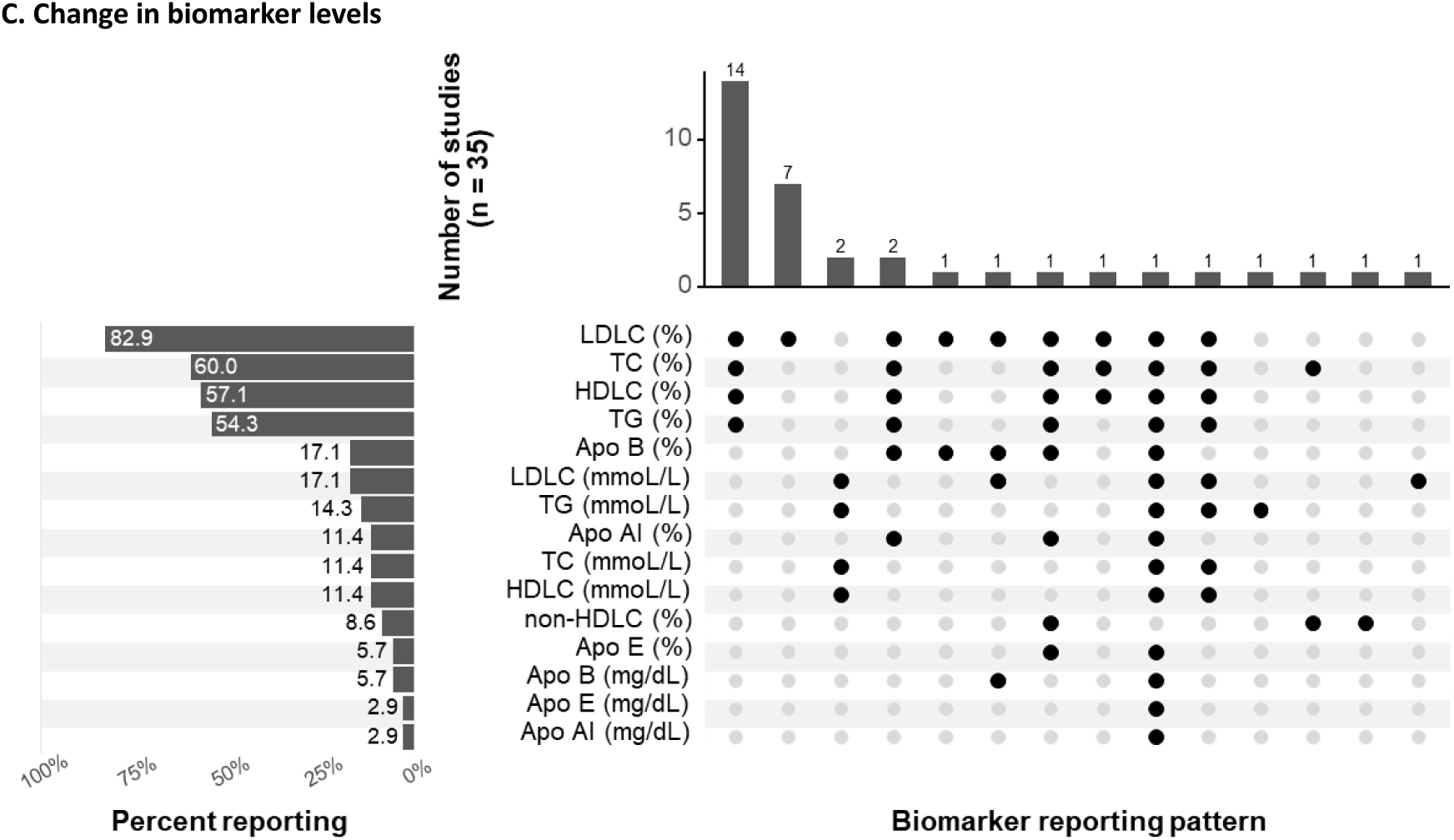
Studies included in the quantitative synthesis (meta-analysis) for the continuous biomarkers. **A.** Studies reporting biomarker measurement before statin treatment. **B.** Studies reporting biomarker measurement after statin treatment. **C.** Studies reporting change in biomarker levels. The left panels show the frequency with which biomarkers were reported. For example, in panel **A**, LDLC (analysed in mmol/L) was the most frequently reported biomarker, appearing in 93.5% (29 out of 31) of the studies that measured biomarkers before statin treatment. On the other hand, the right panels show the reporting patterns. For example, in panel **A**, the most common combination of reported outcomes (first column) included LDLC, TC, TG and HDLC (all analysed in mmol/L), which were reported together in 16 studies. Apo AI = Apolipoprotein AI, Apo B = Apolipoprotein B, Apo E = Apolipoprotein E, HDLC = High-Density Lipoprotein Cholesterol, LDLC = Low-Density Lipoprotein Cholesterol, Lp-a = Lipoprotein(a), TC = Total Cholesterol, TG = Total Triglycerides, VLDLC = Very Low-Density Lipoprotein Cholesterol.

Across all time points, Low-Density Lipoprotein Cholesterol (LDLC) was the most frequently reported continuous biomarker, included in 93.5% of the 31 studies before statin treatment, 100% of the 22 studies after treatment, and 82.9% of the 35 studies reporting biomarker changes (both net/actual and percentage changes). Total Cholesterol (TC) was reported in 77.4% of studies before statin treatment, 86.4% after treatment, and 60.0% of studies reporting biomarker changes. Total Triglycerides (TG) were included in 74.2% of studies before statin treatment, 86.4% after, and 54.3% of studies reporting changes. High-Density Lipoprotein Cholesterol (HDLC) was reported in 74.2% of studies before treatment, 86.4% after, and 57.1% of studies reporting changes. Six other biomarkers were reported in at least two studies but less than a third of studies: Apolipoprotein AI (Apo AI), Apolipoprotein B (Apo B), Apolipoprotein E (Apo E), Lipoprotein(a) [Lp(a)], non-HDLC, and Very Low-Density Lipoprotein Cholesterol (VLDLC), as shown in Figure 2.

### Meta-analysis results

#### Low-Density Lipoprotein Cholesterol (LDLC)

Due to the opposing effects of the *ε2* and *ε4* alleles, individuals with the *ε2ε4* genotype were initially excluded from the analysis. Significant associations between LDLC levels and *APOE* were found for the *ε2* vs. *ε3* (control) genotype both before (22 studies, 4,029 participants; mean difference: −0.31 mmol/L, 95% CI: −0.53 to −0.08, *I²* = 71%) and after statin treatment (17 studies, 2,275 participants; −0.41 mmol/L, 95% CI: −0.68 to −0.14, *I²* = 88%, Tables 1 and S8). For the percentage change in biomarkers before to after statin treatment, *ε2* carriers showed a more pronounced statin effect, with greater reductions in LDLC compared to *ε3* carriers (19 studies, 3,213 participants; mean difference: −2.98%, 95% CI: −5.88% to −0.08%), although heterogeneity was high (*I²* = 81%, Figure 3, Panel A). A sex-stratified analysis partially accounted for this heterogeneity, with the male-specific analysis remaining significant (6 studies, 1,025 participants; mean difference: −5.07%, 95% CI: −7.92% to −2.22%, *I²* = 20%). The analysis of the net/actual change in LDLC levels before and after statin treatment showed a similar pattern to the percentage change, with *ε2* carriers having a greater, although not statistically significant, LDLC reduction (5 studies, 874 participants; mean difference: −0.14 mmol/L, 95% CI −0.28 to 0.003, *I^2^* = 42%, Table 1, Figure S1).

**Figure 3.**
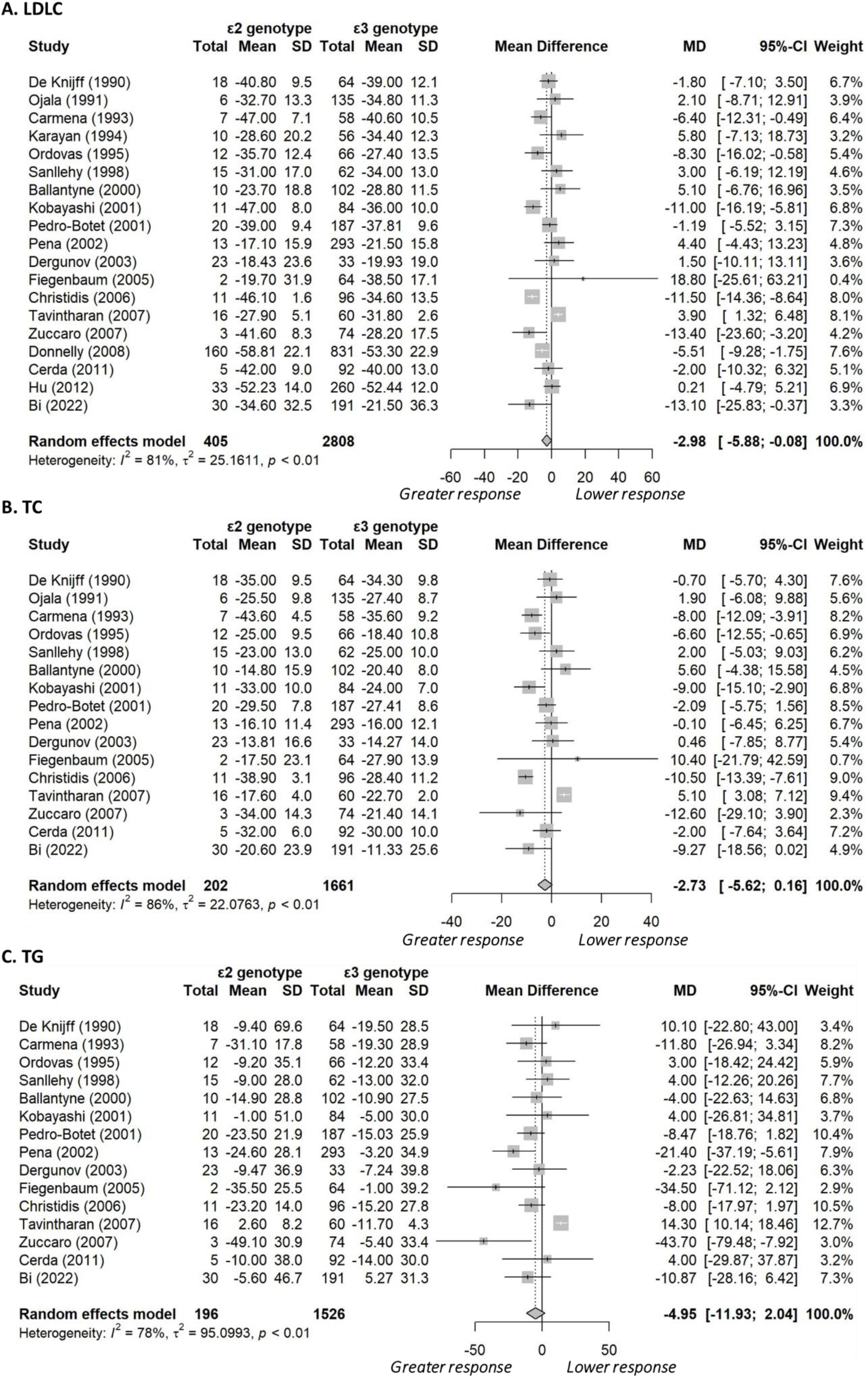
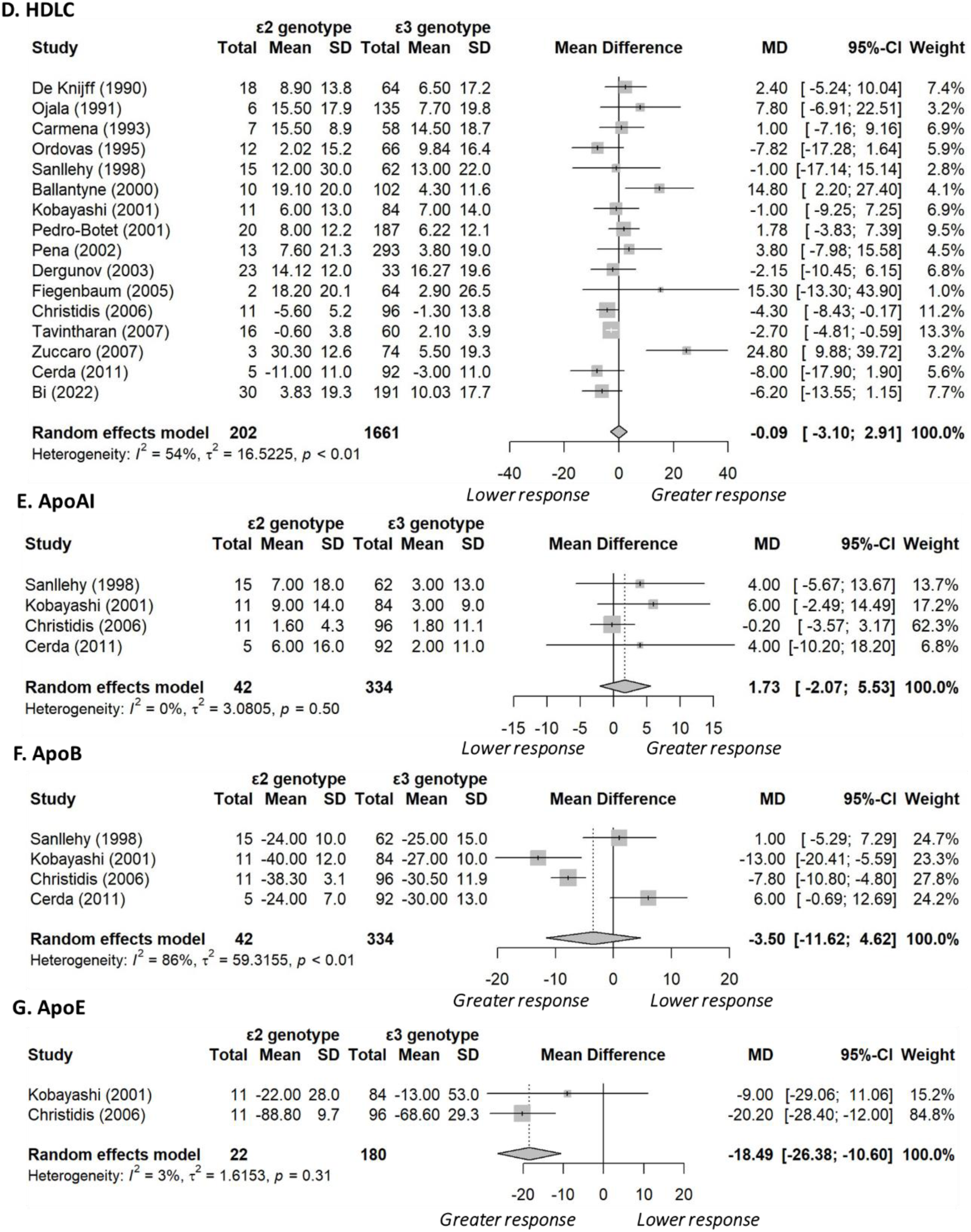
Forest plots comparing Apolipoprotein *ε2* carriers with *ε3* carriers, excluding individuals with the *ε2ε4* genotype. For all biomarkers except HDLC and ApoA, values greater than zero indicate a lower response to statin treatment in *ε2* carriers compared to *ε3* carriers (controls). Abbreviations: Apo AI = Apolipoprotein AI, Apo B = Apolipoprotein B, Apo E = Apolipoprotein E, HDLC = High-Density Lipoprotein Cholesterol, LDLC = Low- Density Lipoprotein Cholesterol, TC = Total Cholesterol, TG = Total Triglycerides.

**Table 1.**
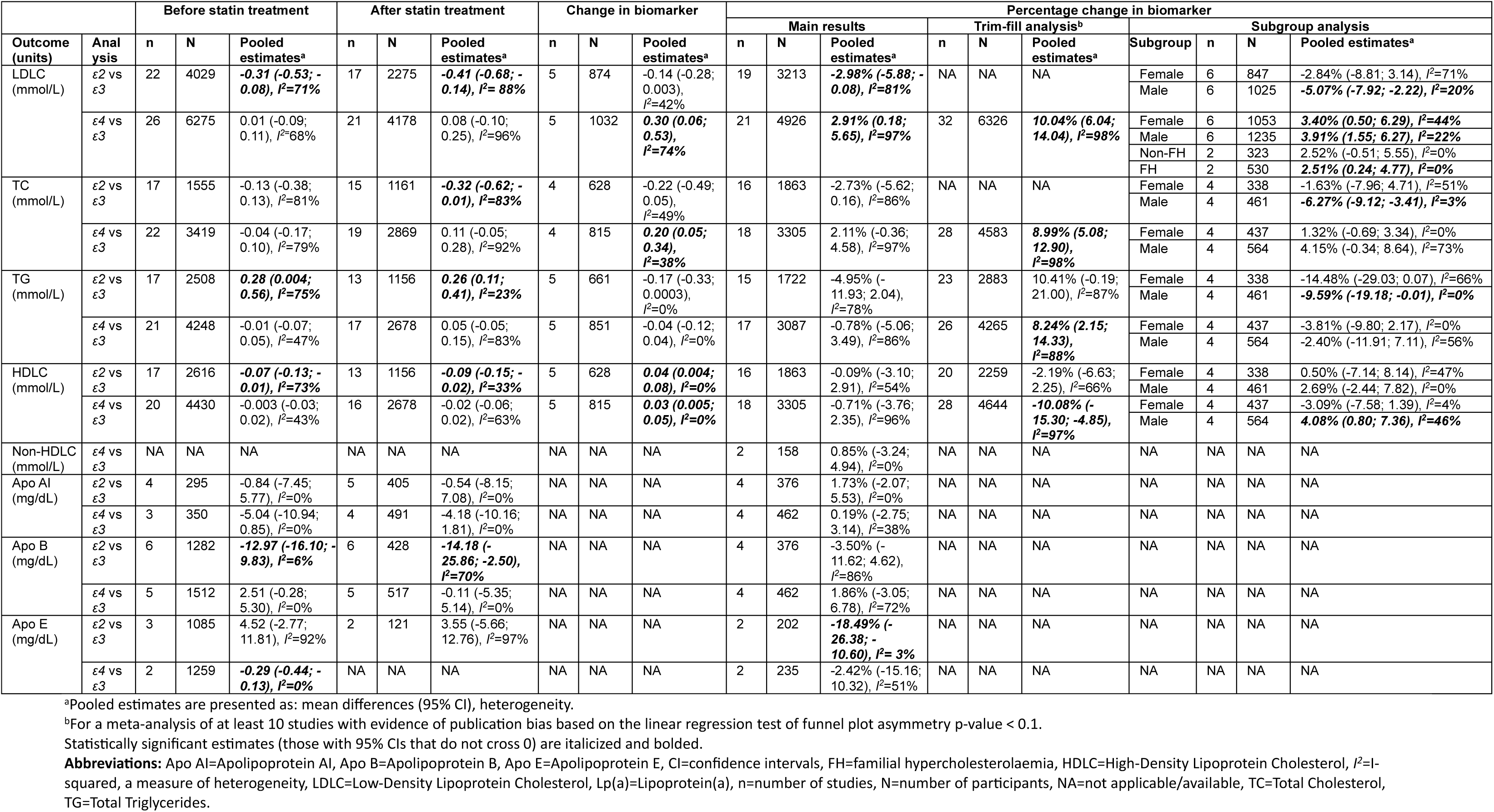
Meta-analysis results with the *ε2ε4* genotype category excluded.

In contrast, LDLC levels for *ε4* vs. *ε3* carriers were similar both before (26 studies, 6,275 participants; mean difference: 0.01 mmol/L, 95% CI: −0.09 to 0.11, *I²* = 68%) and after statin treatment (21 studies, 4,178 participants; mean difference: 0.08 mmol/L, 95% CI: −0.10 to 0.25, *I²* = 96%). *ε4* carriers showed a less pronounced percentage decrease in LDLC compared to *ε3* carriers (21 studies, 4,926 participants; mean difference: 2.91%, 95% CI: 0.18% to 5.65%, Figure 4, Panel A). However, heterogeneity was high (*I²* = 97%), and there was evidence of publication bias (linear regression test of funnel plot asymmetry *P* = 0.002, Figure S2). Sex-stratified analysis explained some of the heterogeneity, with results becoming significant in both females (6 studies, 1,053 participants; mean difference: 3.40%, 95% CI: 0.50% to 6.29%, *I²* = 44%) and males (6 studies, 1,235 participants; mean difference: 3.91%, 95% CI: 1.55% to 6.27%, *I²* = 22%). A subgroup analysis of only Familial Hypercholesterolemia participants (2 studies, 530 participants; mean difference: 2.51%, 95% CI: 0.24% to 4.77%, *I^2^* = 0%) also accounted for the heterogeneity, with the results remaining significant. A trim-and-fill analysis estimated 11 missing studies (32 studies in total, 6326 participants), suggesting that these missing trials would further reduce the LDLC response to statins in *ε4* carriers to 10.04% (95% CI: 6.04% to 14.04%, *I²* = 98%, Figure S2). Consistent with the percentage change, *ε4* carriers had a smaller net reduction in LDLC levels by 0.30 mmol/L (95% CI: 0.06 to 0.53, *I^2^* = 74%; 5 studies, 1,032 participants, Table 1, Figure S3).

**Figure 4.**
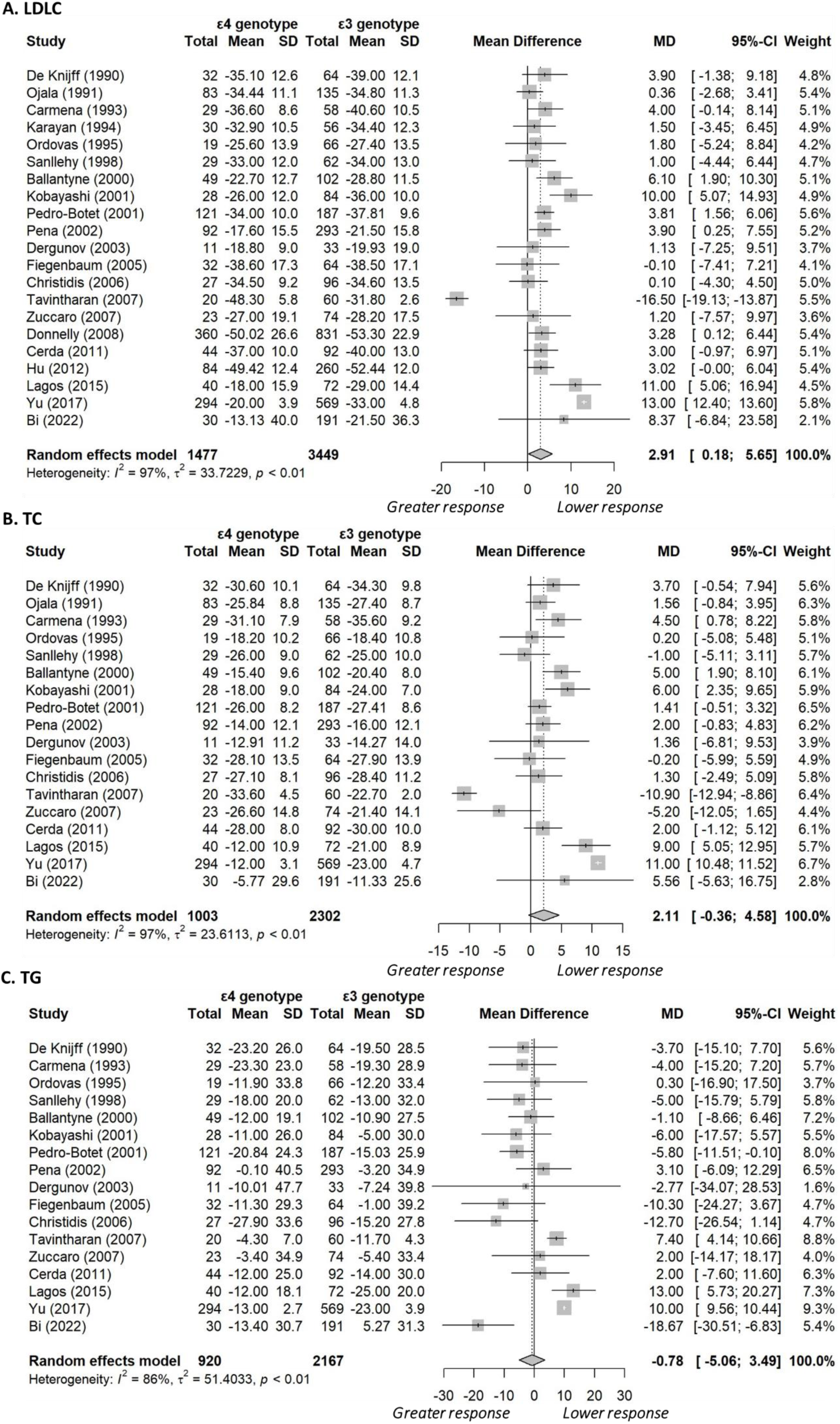
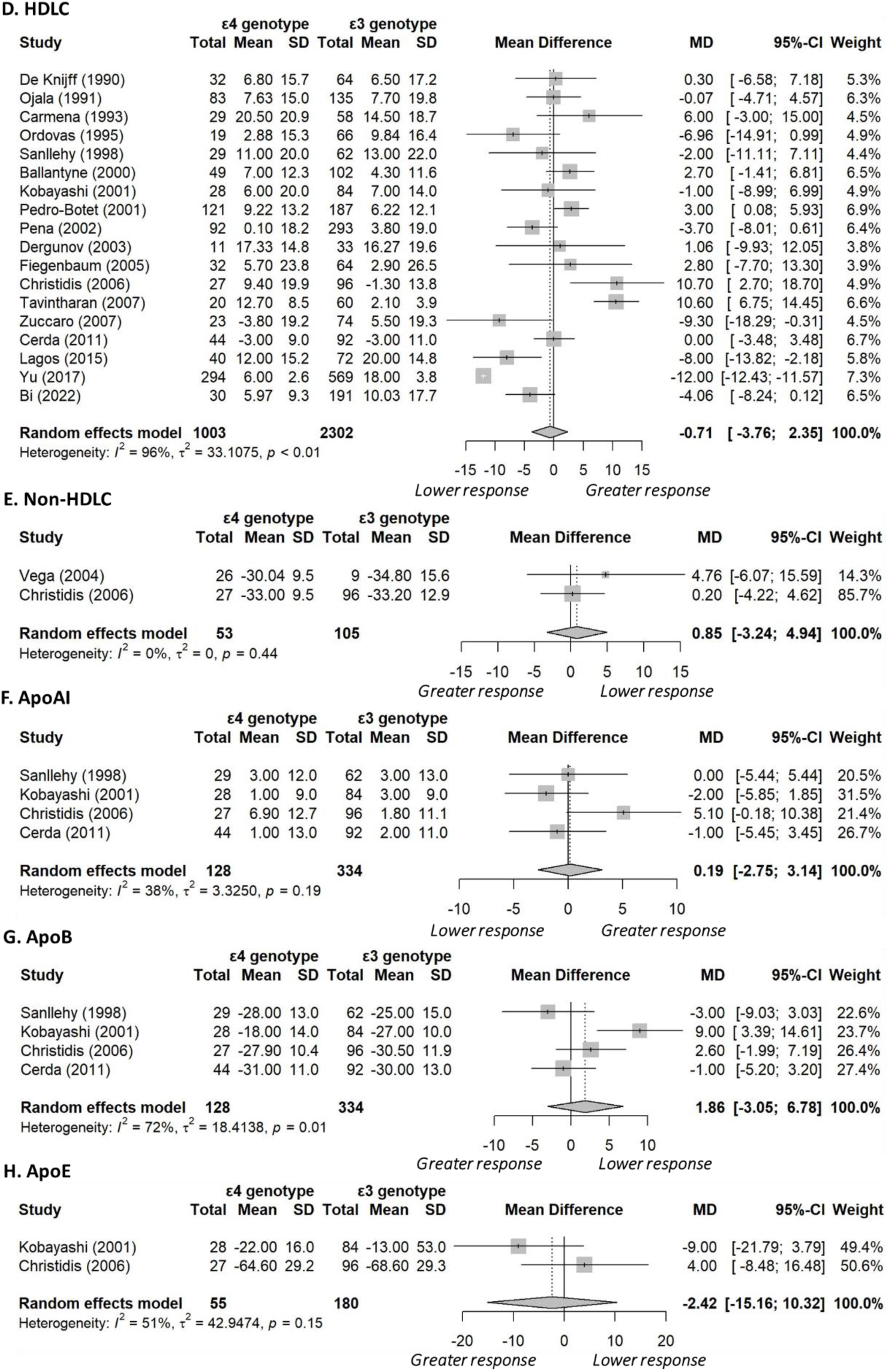
Forest plots comparing Apolipoprotein *ε4* carriers with *ε3* carriers, excluding individuals with the *ε2ε4* genotype. For all biomarkers except HDLC and ApoA, values greater than zero indicate a lower response to statin treatment in *ε4* carriers compared to *ε3* carriers (controls). Abbreviations: Apo AI = Apolipoprotein AI, Apo B = Apolipoprotein B, Apo E = Apolipoprotein E, HDLC = High-Density Lipoprotein Cholesterol, LDLC = Low- Density Lipoprotein Cholesterol, TC = Total Cholesterol, TG = Total Triglycerides.

When individuals with the *ε2ε4* genotype were included in the analysis and categorized under *ε2* carriers (Table S9), *ε3* carriers (Table S10), or *ε4* carriers (Table S11), the results remained consistent with the above results that excluded the *ε2ε4* genotype category. Since the analyses, both including and excluding the *ε2ε4* genotype, showed similar patterns across biomarkers, the subsequent results will focus on analyses that exclude the *ε2ε4* genotype. Additionally, the focus will be on the change in biomarkers. However, full results, including comparisons of *ε2* vs. non-*ε2* and *ε4* vs. non-*ε4* carriers, are available in Tables S8–S11.

#### Total Cholesterol (TC)

No significant associations were found between the percentage (16 studies, 1,863 participants; mean difference: −2.73%, 95% CI: −5.62% to 0.16%, *I²* = 86%, Figure 3, Panel C) and net (4 studies, 628 participants; mean difference: −0.22 mmol/L, 95% CI −0.49 to 0.05, *I²* = 49%, Figure S1) changes in TC levels between the *ε2* and *ε3* carriers. However, in a sex-stratified analysis, male *ε2* carriers showed a greater percentage reduction in TC levels compared to male *ε3* carriers (4 studies, 461 participants; mean difference: −6.27%, 95% CI: −9.12% to −3.41%, *I²* = 3%). For the *ε4* vs. *ε3* comparison, a significant association was observed for the net (4 studies, 815 participants; mean difference: 0.20 mmol/L, 95% CI 0.05 to 0.34, *I²* = 38%, Figure S3) but not percentage change (18 studies, 3,305 participants; mean difference: 2.11%, 95% CI: −0.36% to 4.58%, *I²* = 97%, Figure 4, Panel C). For the percentage change, there was some evidence of publication bias (linear regression test of funnel plot asymmetry *P* = 0.004, Figure S4). A trim-and-fill analysis estimated 10 missing studies (28 studies in total, 4,583 participants), and when these were accounted for, it suggested that *ε4* carriers had a lower TC response to statins by 8.99% (95% CI: 5.08% to 12.90%, *I²* = 98%, Figure S4).

#### Total Triglycerides (TG)

Similar to total cholesterol, no significant associations were found between the percentage (15 studies, 1,722 participants; mean difference: −4.95%, 95% CI: −11.93% to 2.04%, *I²* = 78%, Figure 3, Panel B) and net (5 studies, 661 participants; mean difference: −0.17 mmol/L, 95% CI −0.33 to 0.0003, *I²* = 0%, Figure S1) changes in TG levels between the *ε2* and *ε3* carriers, even after conducting a trim-and-fill analysis to address evidence of publication bias for the percentage change (Figure S5). Similarly, no significant associations were observed for the *ε4* vs. *ε3* comparison (percentage change mean difference [17 studies, 3,087 participants]: −0.78%, 95% CI: −5.06% to 3.49%, *I²* = 86%, Figure 4, Panel B; net change mean difference [5 studies, 851 participants]: −0.04 mmol/L, 95% CI: −0.12 to 0.04, *I²* = 0%, Figure S3). However, there was some evidence of publication bias (linear regression test of funnel plot asymmetry *P* < 0.001 for the percentage change, Figure S6). A trim-and-fill analysis estimated nine missing studies, which, when accounted for, suggested that *ε4* carriers, compared to *ε3* carriers, had a lower TG response to statins by 8.24% (95% CI: 2.15% to 14.33%, *I²* = 88%; 26 studies, 4,265 participants, Figure S6).

#### High-Density Lipoprotein Cholesterol (HDLC)

Based on 16 studies (1,863 participants), *ε2* carriers had a similar percentage change in HDLC levels compared to *ε3* carriers (mean difference: −0.09%, 95% CI: −3.10% to 2.91%, *I²* = 54%, Figure 3, Panel D), even after conducting a trim-and-fill analysis to address evidence of publication bias (Figure S7). However, the net increase in HDLC levels for *ε2* carriers was significantly higher by 0.04 mmol/L (95% CI: 0.004 to 0.08, *I²* = 0%; 5 studies, 628 participants, Figure S1). In a comparable way, *ε4* carriers had a similar percentage change in HDLC levels compared to *ε3* carriers (18 studies, 3,305 participants; mean difference: −0.71%, 95% CI: −3.76% to 2.35%, *I²* = 96%, Figure 4, Panel D) but an unexpectedly higher increase in net HDLC levels (5 studies, 815 participants; mean difference: 0.03 mmol/L, 95% CI 0.005 to 0.05, *I²* = 0%). However, there was evidence of publication bias (linear regression test of funnel plot asymmetry *P* < 0.001 for the percentage change, Figure S8). A trim-and-fill analysis estimated 10 missing studies, which, when accounted for, suggested that *ε4* allele carriers had a lower statin response (smaller increase in HDLC levels) compared to *ε3* carriers (28 studies, 4,644 participants; mean difference: −10.08%, 95% CI: −15.30% to −4.85%, *I²* = 97%, Figure S8). In contrast, male *ε4* carriers had a greater increase in HDLC levels than male *ε3* carriers, with a mean difference of 4.08% (95% CI: 0.80% to 7.36%, *I^2^* = 46%) from 4 studies (564 participants).

#### Other biomarkers

Except for Apolipoprotein E (Apo E), where *ε2* carriers showed a significantly greater reduction in Apo E levels compared to *ε3* carriers (2 studies, 202 participants; mean difference: −18.49%, 95% CI: −26.38% to −10.60%, *I²* = 3%, Figure 3, Panel G), none of the remaining biomarkers listed in Table 1 showed significant percentage differences.

Additional analyses (Figure S9) included: LDLC and rs7412 (CT/TT vs. CC, TT vs. CC, and CT vs. CC), Total Cholesterol and rs7412 (CT/TT vs. CC, and CT vs. CC), Mortality and *ε4* carrier status, Risk of lobar intracranial haemorrhage and *ε2ε4*/*ε4ε4* genotypes. Significant associations were found only for:

- LDLC and rs7412 (TT vs. CC) with a mean difference of −4.58 (95% CI: −7.86 to −1.30, *I²* = 0%) across 2 studies with 2,701 participants.
- Risk of lobar intracranial haemorrhage and *ε2ε4* genotypes (compared to *ε3ε3*), with an odds ratio of 7.60 (95% CI: 4.91 to 11.77, *I²* = 0%) across 2 studies with 1,237 participants, and,
- Risk of lobar intracranial haemorrhage and *ε4ε4* genotypes (compared to *ε3ε3*), with an odds ratio of 6.66 (95% CI: 2.52 to 17.61, *I²* = 0%) across 2 studies with 1,238 participants.

### ASReview Performance

Figure 5 shows the proportion of articles selected by ASReview that matched those included by human reviewers. Out of the 242 studies selected by the human reviewers during abstract/title screening (excluding six additional records identified through reference list searching, Figure 1), ASReview ranked 71 (29%) of these studies among its top 242 selections. This corresponds to a sensitivity of 29.3% (71 out of 242) and a specificity of 95.8% (3,939 out of 4,110), indicating that while ASReview accurately identified most studies not included by human reviewers, it was less successful in identifying those that were included. When the analysis was limited to the 68 studies selected by human reviewers for inclusion in the systematic review, ASReview ranked 26 (38%) of these studies among its top 68 selections, which yields a sensitivity of 38.2% (26 out of 68) and a specificity of 99.0% (4,242 out of 4,284).

**Figure 5.**
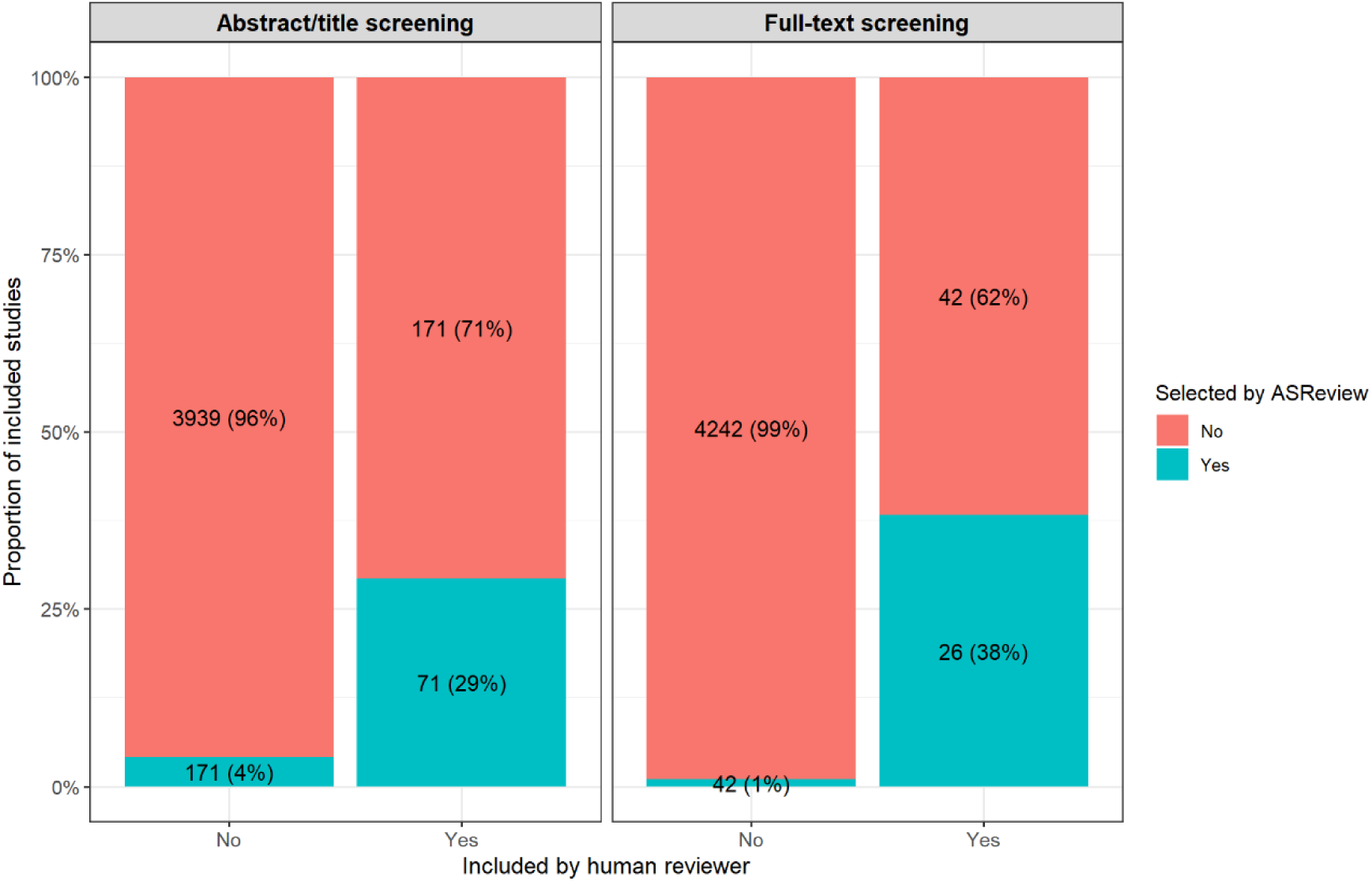
Proportion of articles selected by ASReview that were also included by human reviewers.

## Discussion

We have quantified the association between *APOE* genotype and responses to statins, focusing on the lipid biomarkers, Low-Density Lipoprotein Cholesterol (LDLC), Total Cholesterol (TC), Total Triglycerides (TG), and High-Density Lipoprotein Cholesterol (HDLC). Compared to *ε3* carriers, *ε2* carriers showed a more pronounced response to statins with a greater reduction in LDLC (mean difference: −2.98%, 95% CI: −5.88% to −0.08%), but with similar reductions in TC (−2.73%, −5.62% to 0.16%), and TG (−4.95%, −11.93% to 2.04%), and a similar increase in HDLC (−0.09%, −3.10% to 2.91%). In contrast, after accounting for publication bias, *ε4* carriers, compared with *ε3* carriers experienced smaller reductions in LDLC (10.04%, 6.04% to 14.04%), TC (8.99%, 5.08% to 12.90%), and TG (8.24%, 2.15% to 14.33%), and a smaller increase in HDLC (−10.08%, −15.30% to −4.85%). Previous studies(8, 9, 13, 24–26) have shown some varying results, but in general our findings are consistent with the overall trend of a differential response to statins between *ε2* and *ε4* carriers, when compared with *ε3* carriers. This is also consistent with the biological effects of the *ε2* and *ε4* isoforms in terms of binding to LDL receptors and subsequent downstream effects including clearance.(6, 8, 9)

All *APOE* genotypes derive benefits from statin therapy: the mean percentage reductions in LDLC for *ε2*, *ε3*, and *ε4* carriers were 36.0%, 34.1%, and 31.1%, respectively (Figures 3 and 4, Panel A), which is consistent with the reported mean differences (weighted) of −3.54% (*ε2* vs. *ε3*) and 2.84% (*ε4* vs. *ε3*, before trim-fill analysis). We have focused on percentage changes because net changes do not account for the varying baseline levels across genotypes, and fewer studies reported net changes. Nonetheless, net changes were reported by some studies; for example, based on five studies, the mean difference in LDLC between *ε2* and *ε3* carriers was −0.14 mmol/L (95% CI: −0.28 to 0.003), while for *ε4* versus *ε3*, it was 0.30 mmol/L (95% CI: 0.06 to 0.53) (Table 1 and Figures S1 and S3). All *APOE* genotypes benefited from statin therapy, with the mean net reductions in LDLC levels for *ε2*, *ε3*, and *ε4* carriers being −1.68, −1.52 and −1.23 mmol/L, respectively. It is important to note that these reductions are clinically significant. A prospective meta-analysis involving 90,056 individuals across 14 randomized statin trials found that a 1 mmol/L reduction in LDLC was associated with a 21% reduction in the risk of major coronary events and a 19% reduction in coronary mortality over a 5-year follow-up period.(27) Although *ε4* carriers tend to be more resistant to statin therapy, they may show greater percentage reductions in biomarker levels in response to treatment when their baseline levels are significantly higher (or lower for HDLC) than those of *ε2* or *ε3* carriers, who typically start with lower (or upper for HDLC) baseline levels.(12) As shown in Table 1, baseline biomarker levels for *ε4* and *ε3* genotypes were similar prior to statin treatment, which indicates that the percentage changes observed were probably attributable to statin treatment rather than baseline levels. One potential explanation for the similar baseline levels is that prior non-statin interventions, such as lipid-lowering diets, may mask the effects of *APOE* genotypes on plasma lipid levels.(28, 29)

Heterogeneity was generally high, which is also consistent with previous meta-analyses, and which may be due to factors such as differences in study design, participant characteristics, genotyping procedures, types and doses of statins, duration of treatment, and limited statistical power.(13, 27) In our study, due to limited data, we conducted subgroup analyses based on only sex and Familial Hypercholesterolemia. These analyses were able to account for some or most of the observed heterogeneity. During the sex-stratified analyses, we noticed that males tended to show more pronounced genotype effects. For example, the LDLC analysis comparing *ε2* and *ε3* carriers showed significant effects for males (mean difference: −5.07%, 95% CI: −7.92% to −2.22%) but not females (mean difference: −2.84%, 95% CI −8.81% to 3.14%). These findings are consistent with previous studies, which suggested that these sex differences in response to statin treatment may be linked to variability in immune activation and hormone levels.(30, 31)

To evaluate if a machine learning framework could enhance the efficiency of the screening process, we used ASReview,(15) with stopping criteria based on the number of records selected by human reviewers. While ASReview demonstrated high specificity (95.8% during abstract/title screening and 99.0% during full-text screening), its sensitivity was lower (29.3% for abstract/title screening and 38.2% for full-text screening). This indicates that ASReview effectively identified most studies not included by human reviewers but was less effective at identifying those that were included. In related research, Tran and colleagues recently found that OpenAI’s GPT-3.5 Turbo achieved sensitivities ranging from 81.1% to 96.5% and specificities from 25.8% to 80.4% under the balanced rule, and sensitivities from 94.6% to 99.8% and specificities from 2.2% to 46.6% under the sensitive rule.(32) Thus, GPT-3.5 Turbo, like ASReview, has the potential to reduce the number of citations requiring human screening, although it may miss some citations at the full-text level. As machine learning technology evolves, its performance will improve, and open-source frameworks like ASReview offer opportunities for collaborative enhancements.

In addition to significant heterogeneity, another limitation was the small number of studies (two or fewer) available for certain biomarkers, such as Apo E. Unfortunately, there was a lack of standardized genetic reporting with few studies provided specific genotypes (*ε2ε2*, *ε2ε3*, *ε2ε4*, *ε3ε3*, *ε3ε4*, *ε4ε4*), while others reported only *ε2*, *ε3*, or *ε4* carrier status. We were also unable to account for the specific types and doses of statins used in the included studies, nor could we make dose-equivalence adjustments, which is another key limitation, as not all statins have equal potency. From a clinical perspective, the reduced effect on lipid levels seen in *ε4* carriers could potentially be counteracted by higher (tolerated) doses. We also planned to use the STROPs checklist,(16) but we were unable to apply this tool due to a frequent lack of required information (more than half of the studies were published before 2010 when reporting guidelines specifying mandatory information were not yet available). In future analyses, we plan to overcome some of these limitations by using large more homogeneous populations such as the UK Biobank – such biobanks also have clinical and genetic information at an individual level, which facilitates the analysis of any genetic contrast or analysis of patients with standardized phenotypes. Finally, we did not have sufficient data to report on clinical outcomes such as mortality, but as stated above, a reduction of 1 mmol/L in LDLC is associated with a 19% decrease in the risk of in coronary mortality over a five-year period.(27) Interestingly, our systematic review does show an increased risk of lobar intracranial haemorrhage in *ε4* carriers, with high odds ratios (6.66-7.60). This is not surprising given that *ε4* carriage increases the risk of cerebral amyloid angiopathy and spontaneous intracerebral haemorrhage.

In conclusion, our meta-analysis shows that the *APOE* genotype significantly influences the effectiveness of statin therapy. *ε2* carriers generally show more pronounced reductions in lipid levels, indicating greater responsiveness to treatment, while *ε4* carriers show a comparatively weaker response. These findings should be considered alongside other risk factors, as homozygous *ε2* carriers, in rare cases, may be predisposed to familial dysbetalipoproteinemia, which increases cardiovascular disease risk due to abnormal lipid metabolism.(3, 11) Personalized treatment strategies that consider *APOE* genotype could optimize lipid management and reduce cardiovascular risk across different patient populations. It is important to note that we are not advocating determination of *APOE* genotype prior to statin therapy largely because of the ethical implications, but where the *APOE* genotype is known (and this is likely to increase), it should be considered in terms of both dose and potency of the statin.

## Data Availability

The data underlying this article are available in the article and in its online supplementary material.

