## Supplementary Figures for "APOE genotype and the effect of statins: a systematic review and meta-analysis"

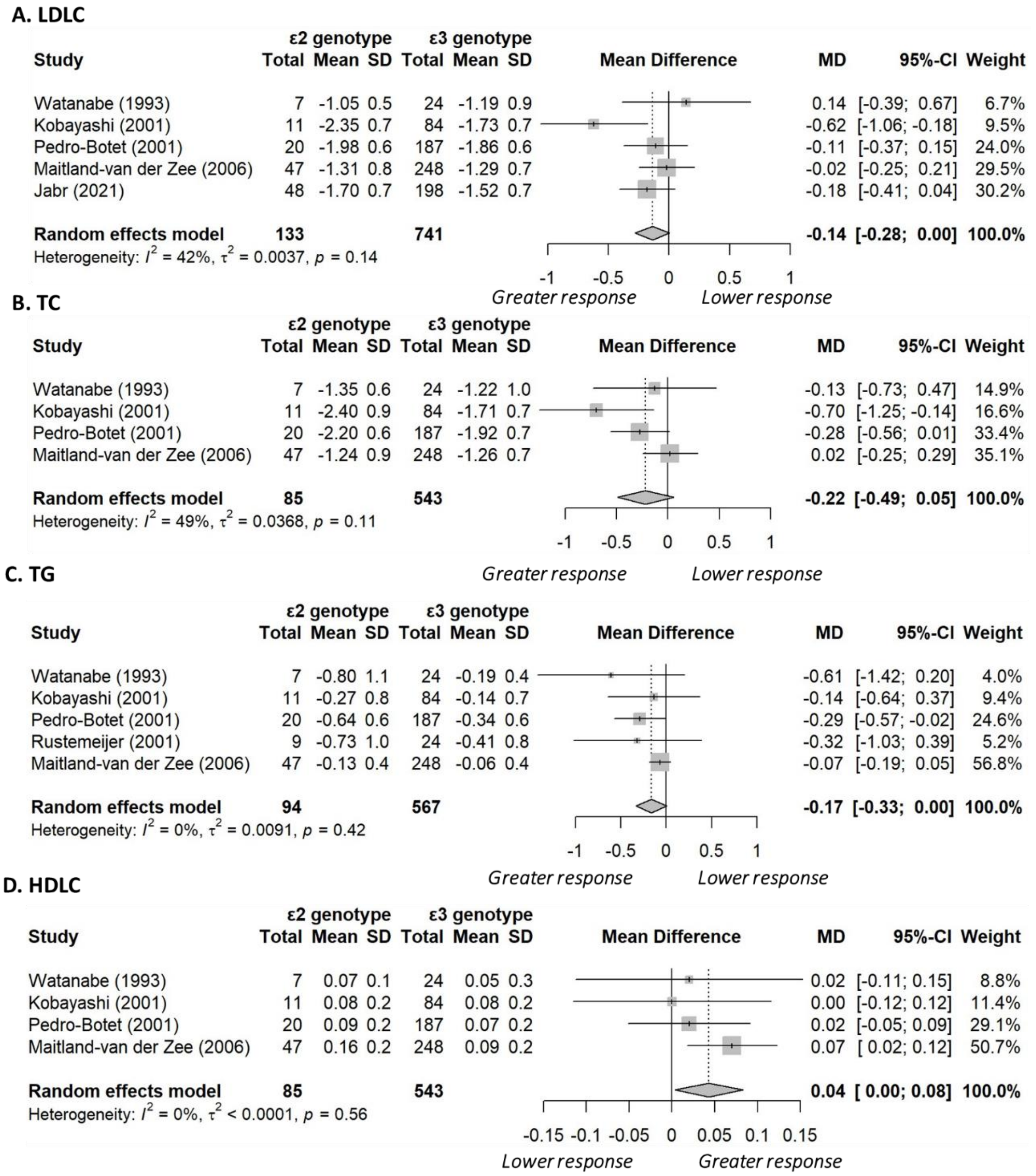


**Figure S1. Forest plots comparing Apolipoprotein *ε2* carriers with *ε3* carriers, excluding individuals with the *ε2ε4* genotype.** All biomarkers are in mmol/L. For all biomarkers except HDLC, values greater than zero indicate a lower response to statin treatment in *ε2* carriers compared to *ε3* carriers (controls). Abbreviations: HDLC = High-Density Lipoprotein Cholesterol, LDLC = Low-Density Lipoprotein Cholesterol, TC = Total Cholesterol, TG = Total Triglycerides.

**A.**

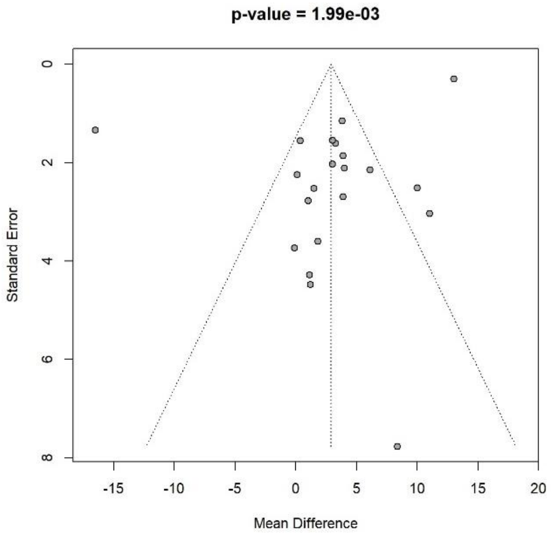


**B.**

| Study | MD | SE(MD) | ε4 Total | ε3 Total | MD | 95%-CI | Weight |
|---|---|---|---|---|---|---|---|
| De Knijff (1990) | 3.9000 | 2.6924 | 32 | 64 | 3.90 | [ -1.38; 9.18] | 3.2% |
| Ojala (1991) | 0.3627 | 1.5547 | 83 | 135 | 0.36 | [ -2.68; 3.41] | 3.3% |
| Carmena (1993) | 4.0000 | 2.1098 | 29 | 58 | 4.00 | [ -0.14; 8.14] | 3.2% |
| Karayan (1994) | 1.5000 | 2.5252 | 30 | 56 | 1.50 | [ -3.45; 6.45] | 3.2% |
| Ordovas (1995) | 1.8000 | 3.5942 | 19 | 66 | 1.80 | [ -5.24; 8.84] | 3.0% |
| Sanllehy (1998) | 1.0000 | 2.7733 | 29 | 62 | 1.00 | [ -4.44; 6.44] | 3.1% |
| Ballantyne (2000) | 6.1000 | 2.1420 | 49 | 102 | 6.10 | [ 1.90; 10.30] | 3.2% |
| Kobayashi (2001) | 10.0000 | 2.5166 | 28 | 84 | 10.00 | [ 5.07; 14.93] | 3.2% |
| Pedro-Botet (2001) | 3.8130 | 1.1470 | 121 | 187 | 3.81 | [ 1.56; 6.06] | 3.3% |
| Pena (2002) | 3.9000 | 1.8610 | 92 | 293 | 3.90 | [ 0.25; 7.55] | 3.2% |
| Dergunov (2003) | 1.1300 | 4.2776 | 11 | 33 | 1.13 | [ -7.25; 9.51] | 2.9% |
| Fiegenbaum (2005) | -0.1000 | 3.7312 | 32 | 64 | -0.10 | [ -7.41; 7.21] | 3.0% |
| Christidis (2006) | 0.1000 | 2.2435 | 27 | 96 | 0.10 | [ -4.30; 4.50] | 3.2% |
| Tavintharan (2007) | -16.5000 | 1.3397 | 20 | 60 | -16.50 | [-19.13; -13.87] | 3.3% |
| Zuccaro (2007) | 1.2000 | 4.4721 | 23 | 74 | 1.20 | [ -7.57; 9.97] | 2.9% |
| Donnelly (2008) | 3.2828 | 1.6120 | 360 | 831 | 3.28 | [ 0.12; 6.44] | 3.3% |
| Cerda (2011) | 3.0000 | 2.0272 | 44 | 92 | 3.00 | [ -0.97; 6.97] | 3.2% |
| Hu (2012) | 3.0200 | 1.5410 | 84 | 260 | 3.02 | [ -0.00; 6.04] | 3.3% |
| Lagos (2015) | 11.0000 | 3.0332 | 40 | 72 | 11.00 | [ 5.06; 16.94] | 3.1% |
| Yu (2017) | 13.0000 | 0.3037 | 294 | 569 | 13.00 | [ 12.40; 13.60] | 3.3% |
| Bi (2022) | 8.3700 | 7.7627 | 30 | 191 | 8.37 | [ -6.84; 23.58] | 2.2% |
| Filled: Hu (2012) | 20.1035 | 1.5410 | 84 | 260 | 20.10 | [ 17.08; 23.12] | 3.3% |
| Filled: Cerda (2011) | 20.1235 | 2.0272 | 44 | 92 | 20.12 | [ 16.15; 24.10] | 3.2% |
| Filled: Ordovas (1995) | 21.3235 | 3.5942 | 19 | 66 | 21.32 | [ 14.28; 28.37] | 3.0% |
| Filled: Karayan (1994) | 21.6235 | 2.5252 | 30 | 56 | 21.62 | [ 16.67; 26.57] | 3.2% |
| Filled: Zuccaro (2007) | 21.9235 | 4.4721 | 23 | 74 | 21.92 | [ 13.16; 30.69] | 2.9% |
| Filled: Dergunov (2003) | 21.9935 | 4.2776 | 11 | 33 | 21.99 | [ 13.61; 30.38] | 2.9% |
| Filled: Sanllehy (1998) | 22.1235 | 2.7733 | 29 | 62 | 22.12 | [ 16.69; 27.56] | 3.1% |
| Filled: Ojala (1991) | 22.7608 | 1.5547 | 83 | 135 | 22.76 | [ 19.71; 25.81] | 3.3% |
| Filled: Christidis (2006) | 23.0235 | 2.2435 | 27 | 96 | 23.02 | [ 18.63; 27.42] | 3.2% |
| Filled: Fiegenbaum (2005) | 23.2235 | 3.7312 | 32 | 64 | 23.22 | [ 15.91; 30.54] | 3.0% |
| Filled: Tavintharan (2007) | 39.6235 | 1.3397 | 20 | 60 | 39.62 | [ 37.00; 42.25] | 3.3% |
| **Random effects model** | | | **1879** | **4447** | **10.04** | **[ 6.04; 14.04]** | **100.0%** |

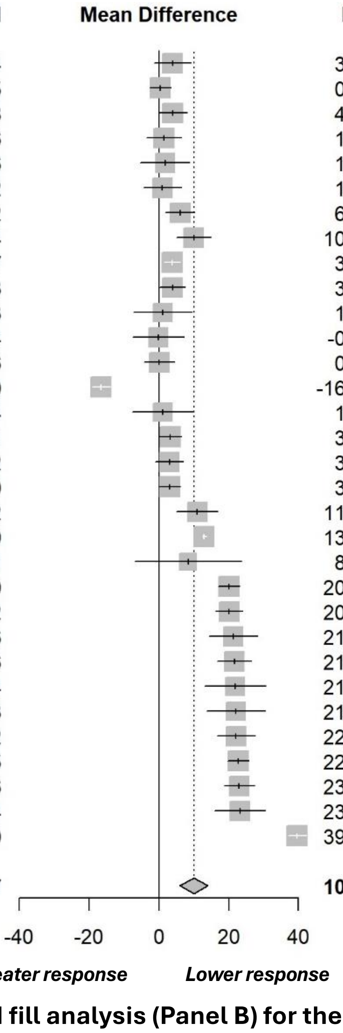


Heterogeneity: $I^2$ = 98%, $\tau^2$ = 124.9740, $p$ < 0.01

**Figure S2. Funnel plot (Panel A) and Trim and fill analysis (Panel B) for the comparison between Low-Density Lipoprotein Cholesterol and Apolipoprotein *ε4* carriers with *ε3* carriers, excluding individuals with the *ε2ε4* genotype.** The p-value for the linear regression test of funnel plot asymmetry is displayed at the top of the figure.

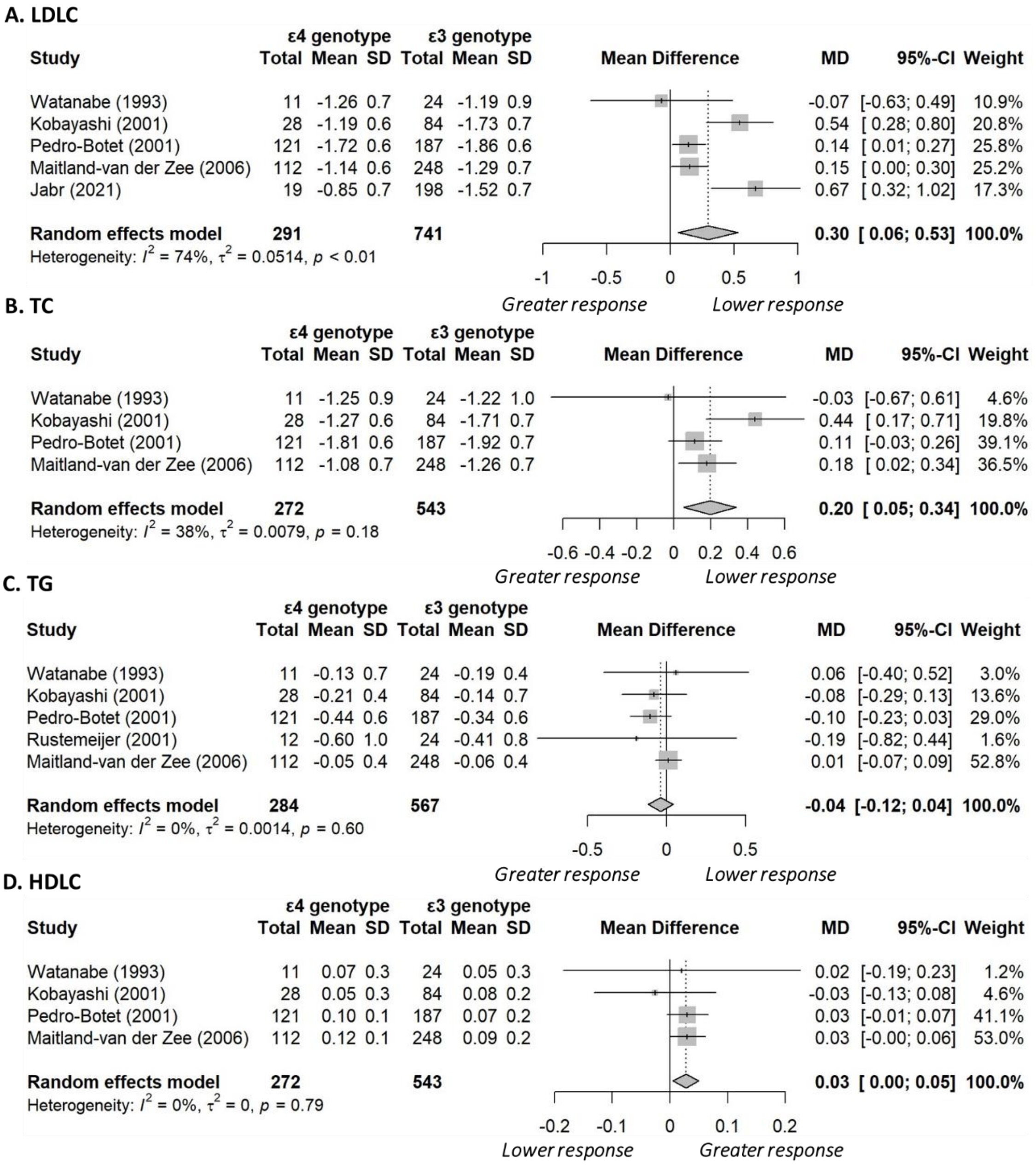


**Figure S3. Forest plots comparing Apolipoprotein *ε4* carriers with *ε3* carriers, excluding individuals with the *ε2ε4* genotype.** All biomarkers are in mmol/L. For all biomarkers except HDLC, values greater than zero indicate a lower response to statin treatment in *ε4* carriers compared to *ε3* carriers (controls). Abbreviations: HDLC = High-Density Lipoprotein Cholesterol, LDLC = Low-Density Lipoprotein Cholesterol, TC = Total Cholesterol, TG = Total Triglycerides.

**A.**

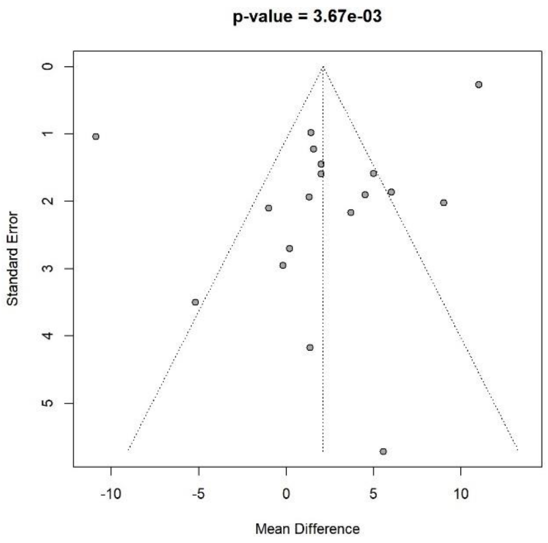


**B.**

| Study | MD | SE(MD) | *ε4* Total | *ε3* Total | MD | 95%-CI | Weight |
|---|---|---|---|---|---|---|---|
| De Knijff (1990) | 3.7000 | 2.1653 | 32 | 64 | 3.70 | [ -0.54; 7.94] | 3.6% |
| Ojala (1991) | 1.5566 | 1.2213 | 83 | 135 | 1.56 | [ -0.84; 3.95] | 3.7% |
| Carmena (1993) | 4.5000 | 1.9004 | 29 | 58 | 4.50 | [ 0.78; 8.22] | 3.6% |
| Ordovas (1995) | 0.2000 | 2.6959 | 19 | 66 | 0.20 | [ -5.08; 5.48] | 3.5% |
| Sanllehy (1998) | -1.0000 | 2.0990 | 29 | 62 | -1.00 | [ -5.11; 3.11] | 3.6% |
| Ballantyne (2000) | 5.0000 | 1.5838 | 49 | 102 | 5.00 | [ 1.90; 8.10] | 3.7% |
| Kobayashi (2001) | 6.0000 | 1.8645 | 28 | 84 | 6.00 | [ 2.35; 9.65] | 3.6% |
| Pedro-Botet (2001) | 1.4060 | 0.9788 | 121 | 187 | 1.41 | [ -0.51; 3.32] | 3.7% |
| Pena (2002) | 2.0000 | 1.4461 | 92 | 293 | 2.00 | [ -0.83; 4.83] | 3.7% |
| Dergunov (2003) | 1.3600 | 4.1690 | 11 | 33 | 1.36 | [ -6.81; 9.53] | 3.2% |
| Fiegenbaum (2005) | -0.2000 | 2.9520 | 32 | 64 | -0.20 | [ -5.99; 5.59] | 3.5% |
| Christidis (2006) | 1.3000 | 1.9330 | 27 | 96 | 1.30 | [ -2.49; 5.09] | 3.6% |
| Tavintharan (2007) | -10.9000 | 1.0388 | 20 | 60 | -10.90 | [-12.94; -8.86] | 3.7% |
| Zuccaro (2007) | -5.2000 | 3.4943 | 23 | 74 | -5.20 | [-12.05; 1.65] | 3.4% |
| Cerda (2011) | 2.0000 | 1.5942 | 44 | 92 | 2.00 | [ -1.12; 5.12] | 3.7% |
| Lagos (2015) | 9.0000 | 2.0175 | 40 | 72 | 9.00 | [ 5.05; 12.95] | 3.6% |
| Yu (2017) | 11.0000 | 0.2674 | 294 | 569 | 11.00 | [ 10.48; 11.52] | 3.8% |
| Bi (2022) | 5.5600 | 5.7099 | 30 | 191 | 5.56 | [ -5.63; 16.75] | 2.9% |
| Filled: Cerda (2011) | 18.4732 | 1.5942 | 44 | 92 | 18.47 | [ 15.35; 21.60] | 3.7% |
| Filled: Ojala (1991) | 18.9166 | 1.2213 | 83 | 135 | 18.92 | [ 16.52; 21.31] | 3.7% |
| Filled: Pedro-Botet (2001) | 19.0672 | 0.9788 | 121 | 187 | 19.07 | [ 17.15; 20.99] | 3.7% |
| Filled: Dergunov (2003) | 19.1132 | 4.1690 | 11 | 33 | 19.11 | [ 10.94; 27.28] | 3.2% |
| Filled: Christidis (2006) | 19.1732 | 1.9330 | 27 | 96 | 19.17 | [ 15.38; 22.96] | 3.6% |
| Filled: Ordovas (1995) | 20.2732 | 2.6959 | 19 | 66 | 20.27 | [ 14.99; 25.56] | 3.5% |
| Filled: Fiegenbaum (2005) | 20.6732 | 2.9520 | 32 | 64 | 20.67 | [ 14.89; 26.46] | 3.5% |
| Filled: Sanllehy (1998) | 21.4732 | 2.0990 | 29 | 62 | 21.47 | [ 17.36; 25.59] | 3.6% |
| Filled: Zuccaro (2007) | 25.6732 | 3.4943 | 23 | 74 | 25.67 | [ 18.82; 32.52] | 3.4% |
| Filled: Tavintharan (2007) | 31.3732 | 1.0388 | 20 | 60 | 31.37 | [ 29.34; 33.41] | 3.7% |
| **Random effects model** | | | **1412** | **3171** | **8.99** | **[ 5.08; 12.90]** | **100.0%** |

Heterogeneity: $I^2 = 98\%$, $\tau^2 = 105.5602$, $p < 0.01$

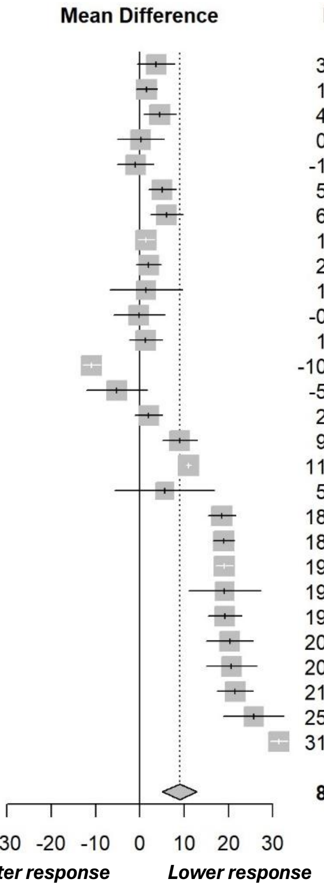


**Figure S4. Funnel plot (Panel A) and Trim and fill analysis (Panel B) for the comparison between Total Cholesterol and Apolipoprotein *ε4* carriers with *ε3* carriers, excluding individuals with the *ε2ε4* genotype.** The p-value for the linear regression test of funnel plot asymmetry is displayed at the top of the figure.

**A.**

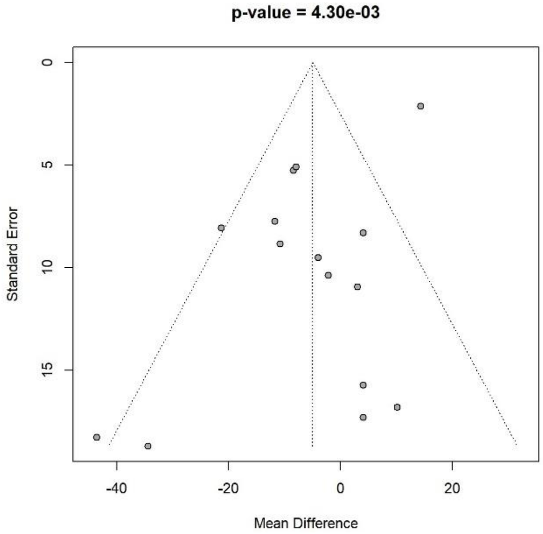


**B.**

| Study | MD | SE(MD) | *ε2* Total | *ε3* Total | MD | 95%-CI | Weight |
|---|---|---|---|---|---|---|---|
| De Knijff (1990) | 10.1000 | 16.7872 | 18 | 64 | 10.10 | [-22.80; 43.00] | 3.5% |
| Carmena (1993) | -11.8000 | 7.7242 | 7 | 58 | -11.80 | [-26.94; 3.34] | 4.8% |
| Ordovas (1995) | 3.0000 | 10.9290 | 12 | 66 | 3.00 | [-18.42; 24.42] | 4.4% |
| Sanllehy (1998) | 4.0000 | 8.2935 | 15 | 62 | 4.00 | [-12.26; 20.26] | 4.7% |
| Ballantyne (2000) | -4.0000 | 9.5057 | 10 | 102 | -4.00 | [-22.63; 14.63] | 4.5% |
| Kobayashi (2001) | 4.0000 | 15.7216 | 11 | 84 | 4.00 | [-26.81; 34.81] | 3.7% |
| Pedro-Botet (2001) | -8.4680 | 5.2515 | 20 | 187 | -8.47 | [-18.76; 1.82] | 5.0% |
| Pena (2002) | -21.4000 | 8.0558 | 13 | 293 | -21.40 | [-37.19; -5.61] | 4.7% |
| Dergunov (2003) | -2.2300 | 10.3527 | 23 | 33 | -2.23 | [-22.52; 18.06] | 4.4% |
| Fiegenbaum (2005) | -34.5000 | 18.6852 | 2 | 64 | -34.50 | [-71.12; 2.12] | 3.2% |
| Christidis (2006) | -8.0000 | 5.0861 | 11 | 96 | -8.00 | [-17.97; 1.97] | 5.1% |
| Tavintharan (2007) | 14.3000 | 2.1238 | 16 | 60 | 14.30 | [10.14; 18.46] | 5.2% |
| Zuccaro (2007) | -43.7000 | 18.2577 | 3 | 74 | -43.70 | [-79.48; -7.92] | 3.3% |
| Cerda (2011) | 4.0000 | 17.2795 | 5 | 92 | 4.00 | [-29.87; 37.87] | 3.4% |
| Bi (2022) | -10.8700 | 8.8236 | 30 | 191 | -10.87 | [-28.16; 6.42] | 4.6% |
| Filled: Ballantyne (2000) | 28.9028 | 9.5057 | 10 | 102 | 28.90 | [10.27; 47.53] | 4.5% |
| Filled: Christidis (2006) | 32.9028 | 5.0861 | 11 | 96 | 32.90 | [22.93; 42.87] | 5.1% |
| Filled: Pedro-Botet (2001) | 33.3708 | 5.2515 | 20 | 187 | 33.37 | [23.08; 43.66] | 5.0% |
| Filled: Bi (2022) | 35.7728 | 8.8236 | 30 | 191 | 35.77 | [18.48; 53.07] | 4.6% |
| Filled: Carmena (1993) | 36.7028 | 7.7242 | 7 | 58 | 36.70 | [21.56; 51.84] | 4.8% |
| Filled: Pena (2002) | 46.3028 | 8.0558 | 13 | 293 | 46.30 | [30.51; 62.09] | 4.7% |
| Filled: Fiegenbaum (2005) | 59.4028 | 18.6852 | 2 | 64 | 59.40 | [22.78; 96.03] | 3.2% |
| Filled: Zuccaro (2007) | 68.6028 | 18.2577 | 3 | 74 | 68.60 | [32.82; 104.39] | 3.3% |
| **Random effects model** | | | **292** | **2591** | **10.41** | **[-0.19; 21.00]** | **100.0%** |

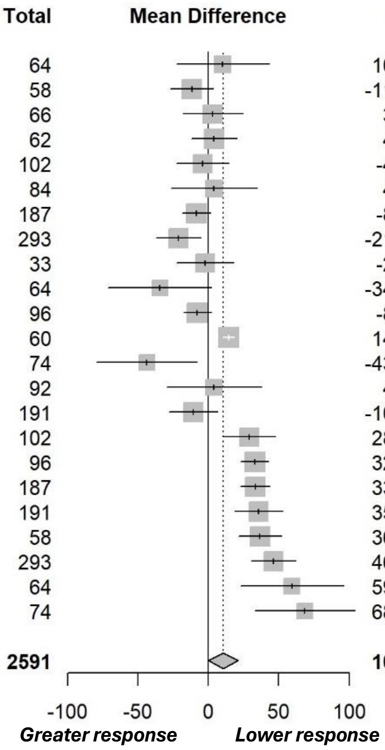


Heterogeneity: $I^2 = 87\%$, $\tau^2 = 552.4668$, $p < 0.01$

**Figure S5. Funnel plot (Panel A) and Trim and fill analysis (Panel B) for the comparison between Total Triglycerides and Apolipoprotein *ε2* carriers with *ε3* carriers, excluding individuals with the *ε2ε4* genotype.** The p-value for the linear regression test of funnel plot asymmetry is displayed at the top of the figure.

**A.**

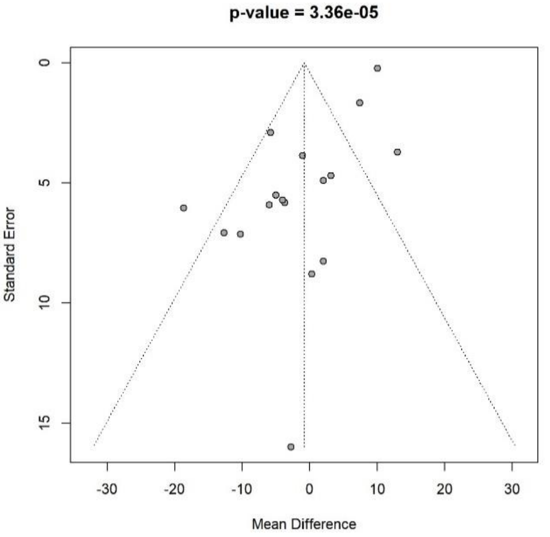


**B.**

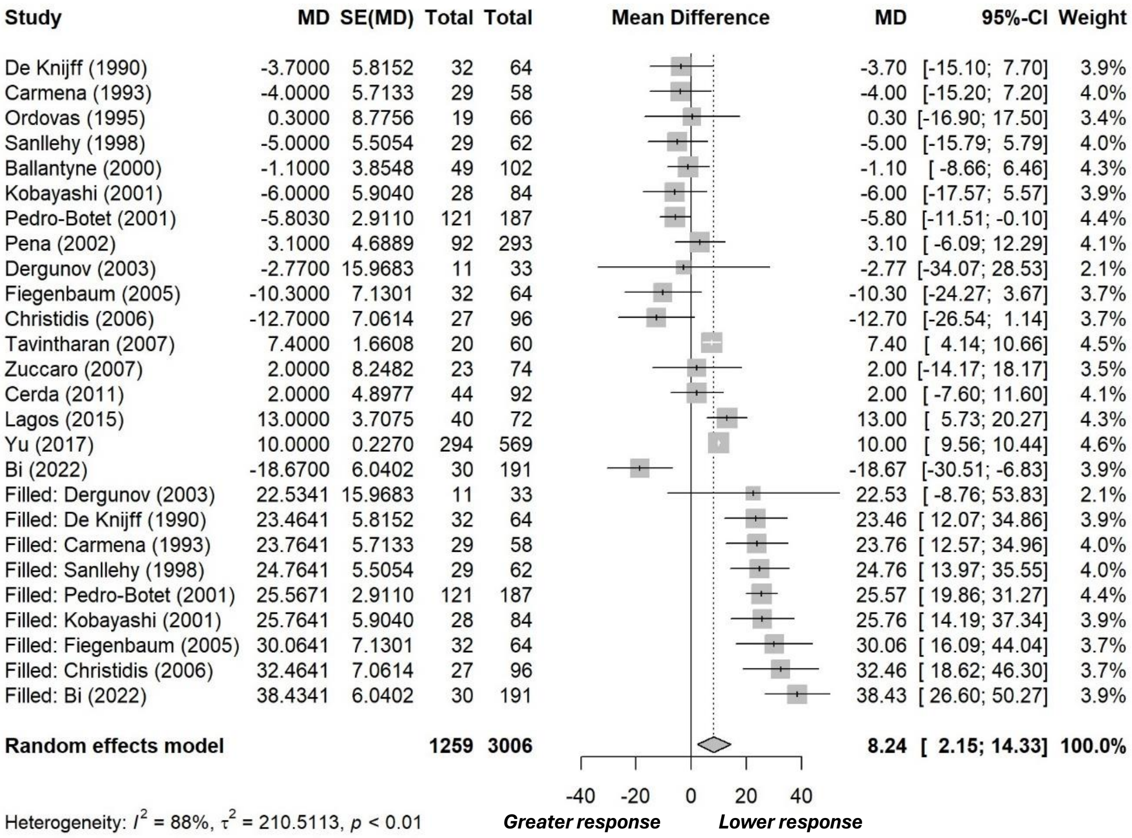


**Figure S6. Funnel plot (Panel A) and Trim and fill analysis (Panel B) for the comparison between Total Triglycerides and Apolipoprotein *ε4* carriers with *ε3* carriers, excluding individuals with the *ε2ε4* genotype.** The p-value for the linear regression test of funnel plot asymmetry is displayed at the top of the figure.

**A.**

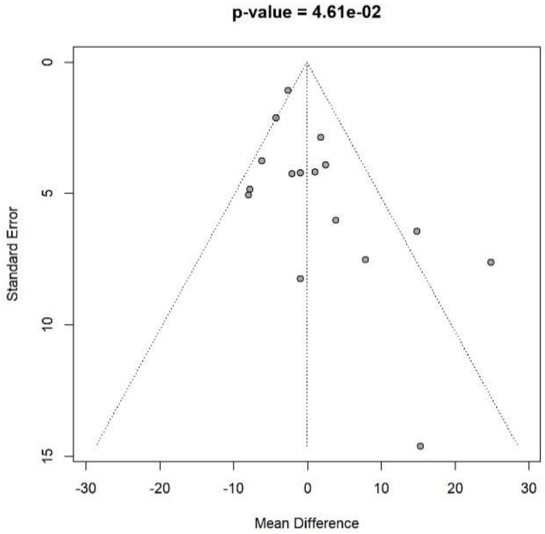


**B.**

| Study | MD | SE(MD) | *ε2* Total | *ε3* Total | MD | 95%-CI | Weight |
|---|---|---|---|---|---|---|---|
| De Knijff (1990) | 2.4000 | 3.8990 | 18 | 64 | 2.40 | [ -5.24; 10.04] | 6.1% |
| Ojala (1991) | 7.8000 | 7.5037 | 6 | 135 | 7.80 | [ -6.91; 22.51] | 4.1% |
| Carmena (1993) | 1.0000 | 4.1647 | 7 | 58 | 1.00 | [ -7.16; 9.16] | 6.0% |
| Ordovas (1995) | -7.8200 | 4.8286 | 12 | 66 | -7.82 | [-17.28; 1.64] | 5.6% |
| Sanllehy (1998) | -1.0000 | 8.2345 | 15 | 62 | -1.00 | [-17.14; 15.14] | 3.8% |
| Ballantyne (2000) | 14.8000 | 6.4280 | 10 | 102 | 14.80 | [ 2.20; 27.40] | 4.7% |
| Kobayashi (2001) | -1.0000 | 4.2068 | 11 | 84 | -1.00 | [ -9.25; 7.25] | 5.9% |
| Pedro-Botet (2001) | 1.7810 | 2.8619 | 20 | 187 | 1.78 | [ -3.83; 7.39] | 6.7% |
| Pena (2002) | 3.8000 | 6.0109 | 13 | 293 | 3.80 | [ -7.98; 15.58] | 4.9% |
| Dergunov (2003) | -2.1500 | 4.2346 | 23 | 33 | -2.15 | [-10.45; 6.15] | 5.9% |
| Fiegenbaum (2005) | 15.3000 | 14.5938 | 2 | 64 | 15.30 | [-13.30; 43.90] | 1.8% |
| Christidis (2006) | -4.3000 | 2.1076 | 11 | 96 | -4.30 | [ -8.43; -0.17] | 7.0% |
| Tavintharan (2007) | -2.7000 | 1.0752 | 16 | 60 | -2.70 | [ -4.81; -0.59] | 7.3% |
| Zuccaro (2007) | 24.8000 | 7.6127 | 3 | 74 | 24.80 | [ 9.88; 39.72] | 4.0% |
| Cerda (2011) | -8.0000 | 5.0513 | 5 | 92 | -8.00 | [-17.90; 1.90] | 5.4% |
| Bi (2022) | -6.2000 | 3.7492 | 30 | 191 | -6.20 | [-13.55; 1.15] | 6.2% |
| Filled: Ojala (1991) | -12.7241 | 7.5037 | 6 | 135 | -12.72 | [-27.43; 1.98] | 4.1% |
| Filled: Ballantyne (2000) | -19.7241 | 6.4280 | 10 | 102 | -19.72 | [-32.32; -7.13] | 4.7% |
| Filled: Fiegenbaum (2005) | -20.2241 | 14.5938 | 2 | 64 | -20.22 | [-48.83; 8.38] | 1.8% |
| Filled: Zuccaro (2007) | -29.7241 | 7.6127 | 3 | 74 | -29.72 | [-44.64; -14.80] | 4.0% |
| **Random effects model** | | | **223** | **2036** | **-2.19** | **[ -6.63; 2.25]** | **100.0%** |

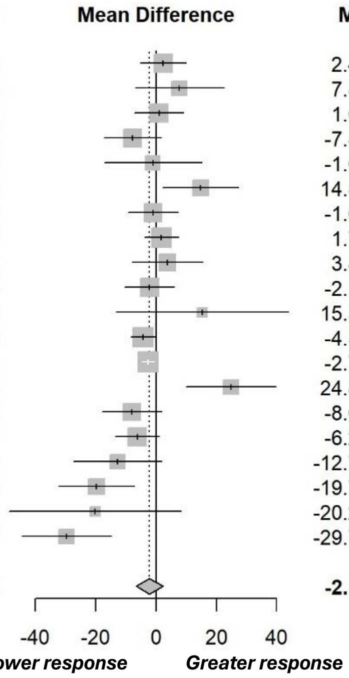


Heterogeneity: $I^2 = 66\%$, $\tau^2 = 68.8415$, $p < 0.01$

**Figure S7. Funnel plot (Panel A) and Trim and fill analysis (Panel B) for the comparison between High-Density Lipoprotein Cholesterol and Apolipoprotein *ε2* carriers with *ε3* carriers, excluding individuals with the *ε2ε4* genotype.** The p-value for the linear regression test of funnel plot asymmetry is displayed at the top of the figure.

**A.**

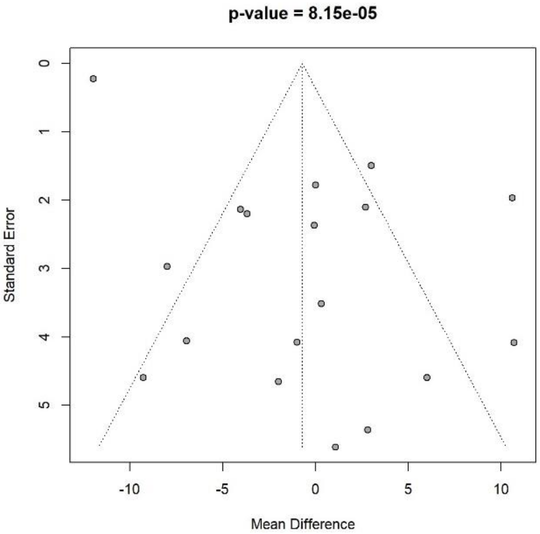


**B.**

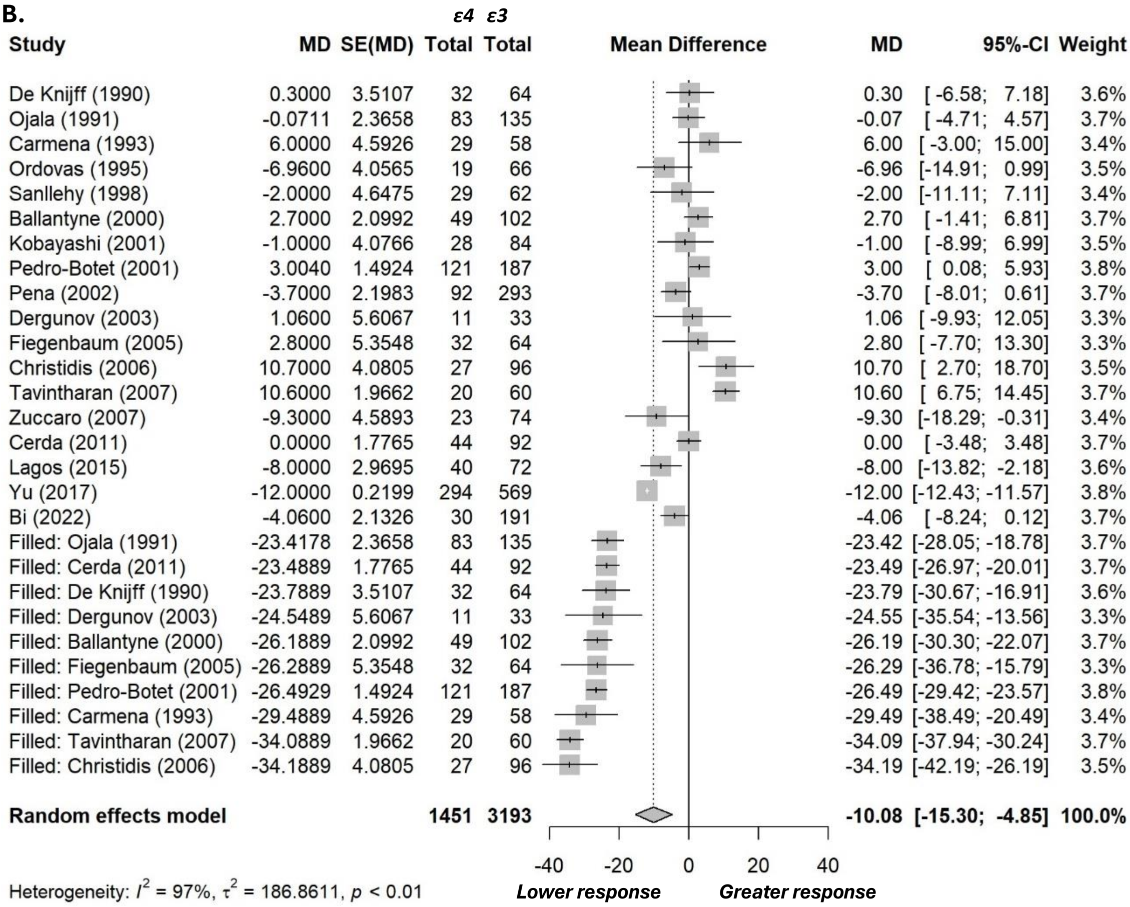


**Figure S8. Funnel plot (Panel A) and Trim and fill analysis (Panel B) for the comparison between High-Density Lipoprotein Cholesterol and Apolipoprotein *ε4* carriers with *ε3* carriers, excluding individuals with the *ε2ε4* genotype.** The p-value for the linear regression test of funnel plot asymmetry is displayed at the top of the figure.

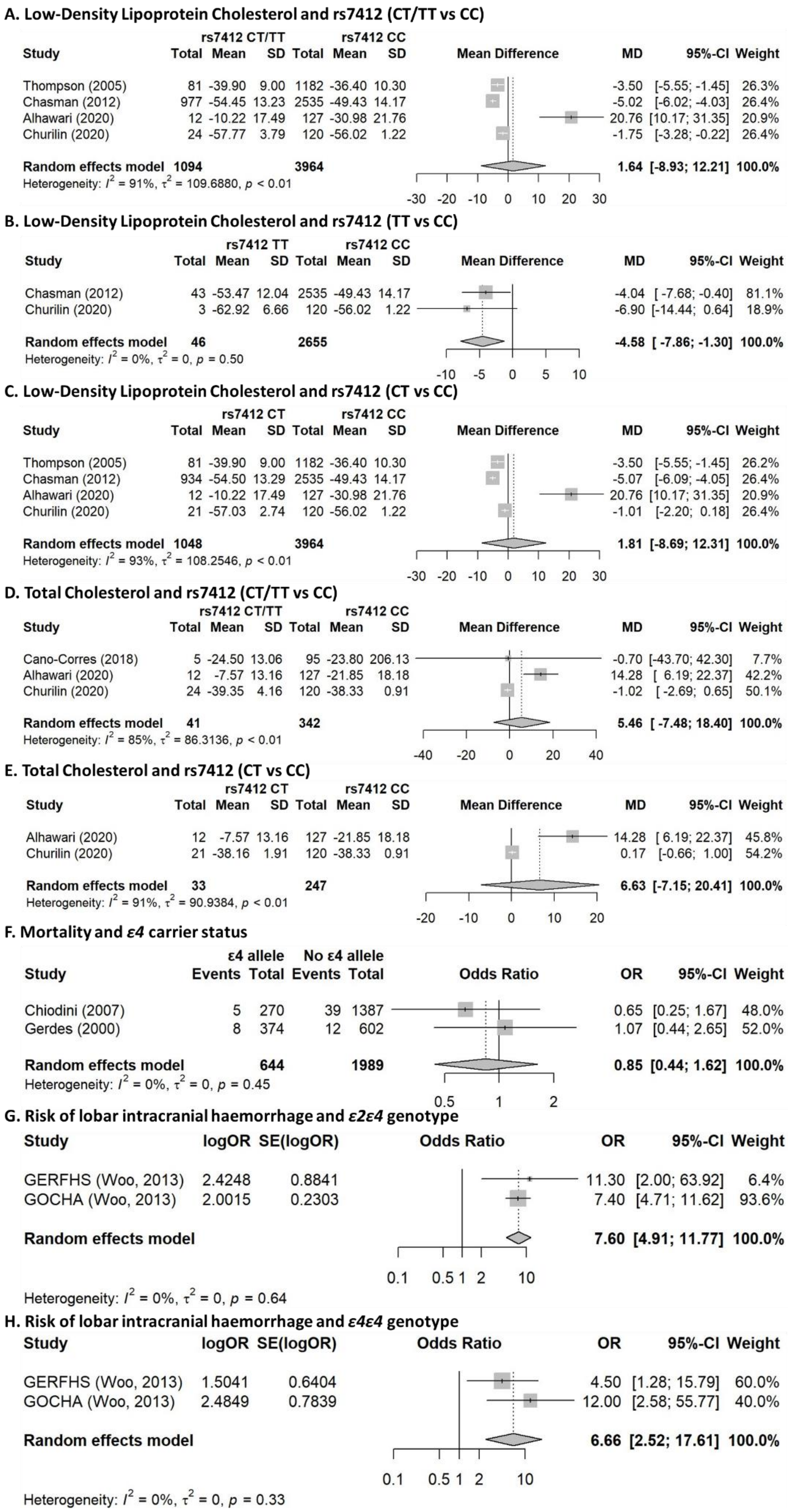


**Figure S9. Forest plots for additional biomarkers.** The reference genotype in panels G and H is *ε3ε3*.
